# A Comparison of Methods for the Optimal Recovery of the Human Fecal Virome

**DOI:** 10.1101/2025.05.12.25327428

**Authors:** Loretta De Chiara, Ryan Doughty, Nuria Estévez-Gómez, Pilar Gallego-García, Pilar Alvariño, Astrid Díez-Martín, Pedro Dávila Piñón, Todd J. Treangen, Joaquín Cubiella, David Posada

**Affiliations:** CINBIO, Universidad de Vigo, Vigo, Spain; Galicia Sur Health Research Institute (IIS Galicia Sur), SERGAS-UVIGO, Vigo, Spain; Department of Computer Science, Rice University, Houston, Texas, USA; Research Group in Gastrointestinal Oncology-Ourense, CIBERehd, Ourense, Spain; Department of Bioengineering, Rice University, Houston, Texas, USA; Ken Kennedy Institute, Rice University, Houston, Texas, USA; Department of Gastroenterology, Health Area of Ourense, Verín and Barco de Valdeorras, Ourense, Spain

**Author notes:** Corresponding authors: Loretta De Chiara and David Posada.

**Keywords:** virome, metagenomics, protocol, human feces

## Abstract

Human virome studies are gaining attention as viruses are increasingly acknowledged as key modulators of microbial communities and human health. However, viral metagenomics presents distinct challenges, including the low abundance and diversity of viruses in biological samples, the lack of universal marker genes, and protocol-induced biases. Although various virome protocols have been benchmarked using viral particles or nucleic acids from mock communities, these often fail to replicate the complexity and heterogeneity of natural viromes. In this study, we systematically evaluated protocol modifications for the metagenomic analysis of human fecal samples, testing alternatives for viral enrichment, nucleic acid extraction, genome amplification, and library preparation. We assessed the impact of each modification on key inferences, including taxonomic and functional assignment, contig quality, viral diversity, and genome structure. Our results highlight important trade-offs between viral genome recovery and contamination removal, underscoring how methodological choices can shape virome composition. Based on our findings, we propose an optimized protocol that enhances the recovery of viral DNA and RNA genomes while minimizing contamination from non-viral sequences, providing a robust framework for future gut virome studies.

## 1. INTRODUCTION

The human gut microbiome is a complex and dynamic ecosystem that plays a crucial role in human health and disease. Numerous studies have established strong associations between the gut microbiome composition and various human conditions [1], including obesity [2], type 2 diabetes [3], cardiovascular disease [4], inflammatory bowel disease [5], colorectal cancer [6], and neurological disorders [7,8]. However, despite increasing knowledge in the field, most studies focus solely on bacteria, overlooking the presence and impact of other microbial components, like viruses, fungi and archaea [9–12].

The viral fraction of the gut microbiome, collectively known as the gut virome, is significantly dominated by prokaryotic viruses (phages) that infect bacteria, representing over 90% of the human gut virome [12,13]. The replication dynamics of phages are tightly linked to bacterial population dynamics, shaping bacterial community structure and composition [14,15]. Phages modulate bacterial metabolism and adaptability either by preventing bacterial adhesion to the mucosa and reducing beneficial commensal bacteria, or by inhibiting the colonization of pathogenic bacteria, thereby promoting host immunity [12,13,16]. Some phages can integrate into their host genomes, transferring new functionalities that can alter bacterial virulence or trigger the development of human immunity [17,18]. Additionally, phages have been suggested to increase intestinal permeability, contributing to a “leaky gut” that facilitates pathogen infiltration and chronic inflammation. In turn, the gut virome is regulated by both bacterial interactions and host immunity.

Viruses are exceptionally difficult to study, as they represent the most genetically diverse microbial group, exhibiting variability in genome type, strandness, genome structure and size, replication strategies, host range, and morphological characteristics. Shotgun metagenomics, which involves sequencing all nucleic acids isolated from a sample, has revolutionized virology by significantly expanding our knowledge of viral diversity [17]. However, detecting viruses using shotgun metagenomics still faces several limitations, including the naturally low-abundance of many viruses, high divergence of virus sequences, and the absence of a universal marker gene [19]. Additionally, bioinformatics methods often struggle due to limitations of alignment-based classification against divergent sequences, and the limited representation of viruses in reference sequence databases [20,21]. As a result, more than 90% of viral sequences in most metagenomic datasets cannot be classified into known viral taxa, a phenomenon known as “viral dark matter” [22–24]. While a significant fraction of this dark matter likely represents genuine novel viruses, sequencing artifacts and laboratory-related issues can also contribute to the unclassified pool of sequences [21,25].

Shotgun metagenomics virome analysis typically involves key steps: purification of viral particles to reduce non-viral contaminants, extraction of viral nucleic acids, amplification of viral genomes, and sequencing library preparation. These steps aim to maximize viral genome recovery while minimizing background noise [11,13]. Most studies use untargeted approaches that enable the detection of both known and novel viruses without prior sequence knowledge. Alternatively, capture-based strategies have been developed to enhance viral detection [26,27], though their reliance on probes designed for known viruses may limit the detection of highly divergent or novel viral sequences.

Laboratory methods can introduce biases in genome representation, significantly influencing virome characterization [28]. Therefore, careful optimization of the methodology is essential for an accurate description of the virome. Different virome protocols have been benchmarked using viral mock communities [29–33]. However, mock communities typically comprise a limited number of viral species, mainly DNA viruses, and fail to replicate the complexity and diversity of viromes in natural samples.

Moreover, many mocks are prepared by pooling extracted viral nucleic acids rather than intact viral particles, and are used at concentrations considerably higher than those encountered in real samples.

In this study, we compared different protocols for sequencing the gut virome in human feces. In particular, we explored the impact of different approaches for (i) viral enrichment, ii) DNA and RNA extraction, iii) genome amplification, and iv) sequencing library preparation. Unlike previous studies that primarily relied on mock viral communities, we used fecal samples, providing insights into protocol performance in a complex, natural virome environment. We assessed the effect of these modifications on key inferences, including taxonomic and functional read assignments, contig characteristics and quality, viral diversity metrics, and genome type and structure classifications. Based on this evaluation, we propose a refined protocol that maximizes the recovery of viral DNA and RNA genomes while minimizing contamination from non-viral sequences, enhancing the detection and characterization of the gut virome in fecal samples.

## 2. METHODS

### 2.1 Study design

We selected a baseline protocol (Protocol A) as the starting point for optimization. This protocol includes virus purification and enrichment, involving sequential centrifugations, filtration, ultracentrifugation, and nuclease treatment (DNase/RNase digestion) to remove unencapsulated free DNA and RNA, followed by RNA extraction and DNA co-purification, retrotranscription of RNA to cDNA, PCR amplification using pseudo-degenerate oligonucleotides (SISPA, Single Primer Amplification), tagmentation-based library preparation, and sequencing (Figure 1).

**Figure 1.**
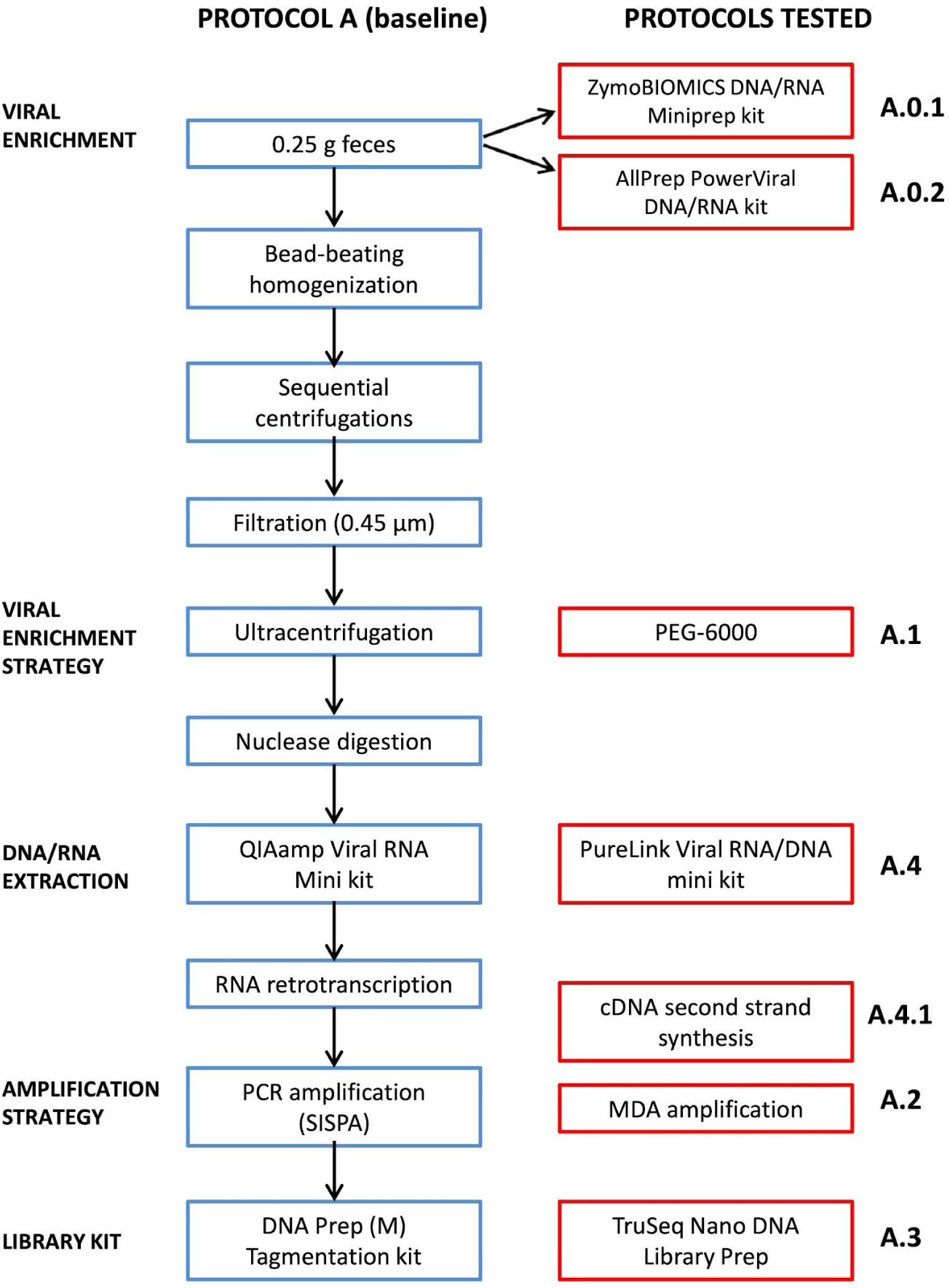
Workflow for shotgun metagenomics sequencing of the fecal virome. The baseline protocol (Protocol A) is shown in blue, while the modifications tested are highlighted in red. Each modification was assessed separately, keeping all other steps consistent with Protocol A. Modifications tested: Protocol A.0.1 and A.0.2: direct nucleic acid extraction without viral enrichment, followed by RNA retrotranscription; Protocol A.1: viral precipitation with PEG-6000 (instead of ultracentrifugation); Protocol A.2: MDA amplification (instead of PCR-SISPA with pseudo-degenerate oligonucleotides); Protocol A.3: library preparation with TruSeq Nano DNA Library Prep (instead of DNA Prep (M) Tagmentation kit); Protocol A.4: DNA/RNA extraction using PureLink Viral RNA/DNA Mini kit (instead of QIAmp Viral RNA Mini kit); Protocol A.4.1: PureLink Viral RNA/DNA Mini kit extraction with second-strand cDNA synthesis (instead of QIAmp Viral RNA Mini kit without second-strand cDNA synthesis).

We systematically evaluated several modifications to key methodological steps in Protocol A (Figure 1):

1. Direct nucleic acid extraction (Protocols A.0.1 and A.0.2): We tested two kits designed to purify nucleic acids for metagenomic studies, avoiding the initial viral enrichment steps.
2. Viral precipitation (Protocol A.1): We carried out polyethylene glycol-based (PEG) precipitation instead of ultracentrifugation.
3. Amplification strategy (Protocol A.2): We performed random amplification using Multiple Displacement Amplification (MDA) instead of PCR-SISPA.
4. Library preparation (Protocol A.3): We used a kit for library construction based on ligation instead of tagmentation.
5. Total viral nucleic acid extraction (Protocol A.4): We tested a kit for extracting both viral DNA and RNA instead of RNA extraction and DNA co-purification.
6. cDNA second-strand synthesis (Protocol A.4.1): We used the extraction kit from Protocol A.4 and tested the addition of a second-strand synthesis step for cDNA.

### 2.2 Samples

Feces samples were collected from 18 patients who had not used antibiotics for at least two months before sampling. Individuals were recruited from Complexo Hospitalario Universitario de Ourense, and the samples were obtained from the Biobank of the Galicia Sur Health Research Institute (IISGS). Sample transfer was approved by the Ethics Committee for Clinical Research of Galicia (2021/134).

Written informed consent was obtained from all participants, ensuring anonymity and adherence to ethical and clinical guidelines established by the Spanish Government and the Declaration of Helsinki.

The 18 samples were distributed as follows: n=5 for protocols A, A.0.1 and A.0.2 (3 men, 2 women), n=2 for protocol A, A.1 and A.2 (2 men), n=5 for protocol A and A.3 (2 men, 3 women), and n=6 for protocols A, A.4 and A.4.1 (4 men, 2 women).

### 2.3 Homogenization

Feces samples were thawed and homogenized using a sterile loop until a uniform consistency was achieved. For all the modifications we used 0.25 g of feces, which was weighed directly into ZR BashingBeads Lysis Tubes containing 0.1 and 0.5 mm beads (Zymo Research, CA, USA). To preserve the nucleic acids we added 750 µL of DNA/RNA Shield (Zymo Research, CA, USA) and performed bead-beating for 10 min in a Vortex-Genie 2 (Scientific Industries, NY, USA) to ensure thorough cell disruption and viral particle release.

### 2.4 Enrichment and precipitation of viral particles

Homogenized fecal samples were centrifuged at 14000 g for 30 s at 8°C, and 400 µL of supernatant was carefully recovered. We added 800 µL of Hanks′ Balanced Salt Solution (HBSS; Sigma, MA, USA) and performed three sequential centrifugation steps (10000 g, 2 min, 8°C) to sediment large fecal aggregates. One mL of the clarified supernatant was finally recovered and filtered through a 0.45 µm pore syringe filter. Viral particles were concentrated using ultracentrifugation (Protocol A) or PEG-based precipitation (Protocol A.1).

For the ultracentrifugation approach (Protocol A), the filtered fecal suspension was transferred to a 6 mL Quick-Seal® Bell-Top Polypropylene Tube (Beckman Coulter Life Sciences, CA, USA) and volume was completed with HBSS. Tubes were heat-sealed and centrifuged at 750000 g for 1 h at 8°C in an Optima™ XPN-100 ultracentrifuge using a 100 Ti fixed-angle rotor (Beckman Coulter, Life Sciences, CA, USA). The supernatant was completely removed without disturbing the pellet containing the viral particles, which was then carefully resuspended in 500 µL of HBSS. The resuspended pellet was stored at-80°C.

For the viral precipitation based on PEG (Protocol A.1), we added a solution of 5 M NaCl molecular biology grade (Promega, Madison, WI, USA) to the filtered fecal suspension for a final concentration ∼1 M. Then, we added PEG-6000 molecular biology grade (Calbiochem, CA, USA) to a final concentration of 10% (w/v), and the tube was inverted several times to mix. Samples were incubated overnight at 4°C to allow viral precipitation. The next day, we centrifuged the tubes at 1200 g for 20 min at 4°C to pellet the PEG-virus complex. The pellet was resuspended in 500 µL of HBSS and stored at-80°C.

The viral enrichment was completed with the digestion of unencapsulated and unenveloped free DNA and RNA. One hundred twenty microliters of the fraction of precipitated viruses were treated with a mixture containing 4.8 U Turbo DNAse and 1X Turbo DNAse buffer (Invitrogen, MA, USA), RNAse cocktail enzyme mix (Invitrogen, MA, USA) containing 4 U RNAse A and 160 U RNAse T1, and 25 U benzonase nuclease, purity >90% (Millipore, MA, USA). This mixture was digested for 1.5 h at 37°C.

### 2.5 Nucleic acid extraction

We extracted viral DNA and RNA from the nuclease-treated product using either QIAamp Viral RNA Mini kit (Qiagen, Hilden, Germany; Protocol A) or the PureLink Viral RNA/DNA Mini kit (Invitrogen, Waltham, MA, USA; Protocol A.4 and A.4.1). Extractions were performed according to the manufacturer’s instructions, but without carrier RNA. Nucleic acids were eluted in 50 µL nuclease-free water (NFW) and stored at-80°C.

For Protocols A.0.1 and A.0.2, we tested two additional extraction kits designed for metagenomic DNA and RNA purification: ZymoBIOMICS DNA/RNA Miniprep kit (Zymo Research, Irvine, CA, USA) and AllPrep PowerViral DNA/RNA kit (Qiagen, Hilden, Germany), respectively. The input sample for both kits was 0.25 g of feces, and extractions were performed following the manufacturers’ recommendations, with minor modifications. Bead-beating was conducted using the beads supplied with each kit. For the ZymoBIOMICS kit, we followed the total nucleic acid purification protocol, while for the AllPrep PowerViral kit we included β-mercaptoethanol to ensure the efficient extraction of RNA viruses. Nucleic acids were eluted in 50 µL NFW and stored at-80°C.

### 2.6 Retrotranscription of RNA viruses and cDNA second-strand synthesis

We retrotranscribed RNA to cDNA using 11 µL of extracted nucleic acid as input, following the SuperScript IV reverse transcriptase (Invitrogen, MA, USA) protocol. The reaction mix included 1 µL of 50 µM pseudo-degenerate oligonucleotides D2_8N (5’-AAGCTAAGACGGCGGTTCGGNNNNNNNN-3’) [34] containing 8 random nucleotides at the 3′ end for random priming and 20 nucleotides of defined sequence at the 5′ end, in a final reaction volume of 20 µL.

For Protocol A.4.1 we used the retrotranscription product (cDNA) as a template for double-stranded cDNA (ds cDNA) synthesis using the Second Strand cDNA Synthesis kit (Invitrogen, MA, USA) following the manufacturers’ recommendations. Residual RNA was removed using RNAse I, and the resulting ds cDNA was purified with the QIAquick PCR Purification Kit (Qiagen, Hilden, Germany).

### 2.7 Genomic amplification

We tested two different genomic amplification strategies: PCR-SISPA (Protocol A) and MDA (Protocol A.2). We used 8 µL of the reverse transcription product (or the second-strand cDNA synthesis product for Protocol A.4.1) as input for PCR-SISPA amplification (Protocol A). The reaction was carried out using Q5 High-Fidelity DNA polymerase (New England Biolabs, MA, USA) in a final volume of 50 µL, containing 3 µL of 20 µM primer D2 (5’-AAGCTAAGACGGCGGTTCGG-3’) [34]. Cycling conditions were: 98°C 1 min, 30 cycles of (98°C 10 s, 55°C 30 s, 72°C 45 s), 72°C 2 min, cool down to 4°C.

For MDA (Protocol A.2), we used the illustra GenomiPhi v2 DNA Amplification kit (GE Healthcare, IL, USA), and amplified 1 µL of the reverse-transcribed product following the manufacturer’s instructions. For each sample, triplicate reactions were performed and subsequently pooled to minimize amplification biases.

Amplification products from both PCR-SISPA and MDA were quantified using the Qubit 3.0 with the dsDNA BR kit (Thermo Fisher Scientific, MA, USA). Their size was checked using the 2200 TapeStation D1000 kit (Agilent Technologies, CA, USA).

### 2.8 Library preparation and sequencing

We generated sequencing libraries from the amplified fragments from Protocol A using two different Illumina library preparation kits using tagmentation (Protocol A) or ligation (Protocol A.3). For Protocol A, libraries were prepared with the DNA Prep (M) Tagmentation kit (Illumina, CA, USA) using ¼ of the recommended reaction volume. A total of ∼125 ng of DNA was used as input and 5 cycles of tagmented DNA amplification were performed. Indexing was done using Nextera™ DNA CD Indexes (Illumina, CA, USA).

For Protocol A.3, libraries were built using the TruSeq Nano DNA Library Prep (Illumina, CA, USA) with 100 ng of DNA input. To avoid excessive fragmentation we modified the protocol by skipping the DNA fragmentation, end-repair and size-selection steps, starting directly from the 3’ adenylation step. Indexing was performed using TruSeq DNA CD Indexes (Illumina, CA, USA).

Finally, for both library methods, we checked the size and quantified the libraries as described above. We sequenced libraries on an Illumina MiniSeq at the sequencing facility of the University of Vigo using the MiniSeq high-output kit (PE150 reads for DNA Prep libraries and SE150 reads for TruSeq libraries; total sequencing output of 7.5 Gb per run). We performed five sequencing runs, multiplexing samples as follows: 17-plex for Protocols A, A.0.1 and A.0.2; 90-plex for Protocols A, A.1 and A.2; 6-plex (in two separate runs) for Protocols A and A.3; 25-plex for Protocol A, A.4 and A.4.1.

### 2.9 Bioinformatics quality control and preprocessing

Raw sequencing reads were downloaded and quality control (e.g. length and low-complexity filtering, adapter removal) was performed using fastp [35] with the following parameters (-l 50-y 3-W 4-M 20

-x). Next, we aligned all reads to the GRCh38 (hg38) human reference genome with Bowtie2 [36] and removed reads that aligned. The quality of reads was assessed before and after each step using FastQC [37] and MultiQC [38]. To adjust for differences in sequencing output (paired vs single-end reads), commands were adjusted accordingly.

### 2.10 Taxonomic and functional assignment

SeqScreen [39] was used in default mode to provide taxonomic classification and functional annotation to each read in every sample. SeqScreen was selected due to its sensitive taxonomic classification, which integrates outputs from Centrifuge [40] and Diamond [41], while also providing functional annotations specific to microbial pathogenesis. Centrifuge uses a RefSeq-based database that includes all archaeal, bacterial, and viral sequences, while Diamond utilizes a curated version of UniRef100 with a focus on microbial pathogens. In cases where Seqscreen provided more than one taxonomic ID for a read, the lowest common ancestor (LCA) of the IDs was taken using Taxonkit [42] and used as the final assignment for that read. If taxonomic assignments for paired-end reads did not match, the LCA was assessed again. Full taxonomic lineages were retrieved from the final taxonomic ID using Taxonkit, enabling higher-order taxonomic comparisons.

The overall percentage of reads assigned by SeqScreen was computed by dividing the number of reads assigned to any taxonomy other than “root” (i.e., the top-level node in the taxonomic tree indicating unclassified reads), by the total number of reads after preprocessing. The percentage of viral and bacterial reads was calculated by dividing the number of reads assigned to viruses or bacteria, respectively, by the number of assigned reads. The virus-to-bacteria ratio was calculated as the number of viral reads divided by the number of bacterial reads. As an additional virome enrichment assessment, we used ViromeQC [43]. This tool calculates three enrichment scores based on reads mapping to the small and large ribosomal RNA subunits (SSU and LSU, respectively), and single-copy bacterial markers. Log2 fold-change values were computed to compare each tested protocol relative to the baseline protocol (Protocol A).

Genome composition and structure were assigned to reads by linking the taxonomic family of each read to reference information from the Virus Metadata Resource (VMR_MSL39_v1, [44]) and ViralZone (SIB Swiss Institute of Bioinformatics, [45]). Higher-order nucleic acid compositions (ssDNA, ssRNA, dsDNA, dsRNA) were calculated by summing all viruses from each type, including positive-and negative-strand forms as well as reverse-transcribing viruses.

### 2.11 Metagenomic assembly and identification of viral sequences

We assembled metagenomic contigs for each sample using MEGAHIT [46] with default parameters. Contigs shorter than 1,000 base pairs (bp) were filtered out to retain only longer sequences for further analysis. All assembled contigs were analyzed using CheckV [47] to identify potential viral contigs, estimate genome completeness and categorize sequences based on their quality.

### 2.12 Richness and diversity metrics

To evaluate the viral diversity profiles of each protocol, we used taxonomic assignments from Seqscreen and calculated common diversity metrics at increasing read cutoffs. For each sample, we filtered viral assignments with at least *n* reads and calculated the number of viral taxa (richness) as well as Shannon [48] and Simpson [49] diversity. The read cutoff (*n*) was progressively increased from 0 to 1000 in steps of 5. Within each protocol, we computed the mean diversity and standard error at each cutoff. This process was performed separately for viruses at both the family and species levels.

This approach enables us to visualize how diversity changes across filtering thresholds, which are commonly used in taxonomic classification studies to remove low-frequency false positives. Additionally, it reveals the relative contribution of rare (low-read) versus abundant (high-read) taxa within each protocol, providing deeper insights into the diversity composition and the impact of read abundance on detected viral profiles.

### 2.13 Statistical analyses

To assess the statistical significance of the different metrics, we applied a Mann-Whitney U-test comparing each protocol against Protocol A. For viral diversity, we assessed overall differences across read cutoffs using the approach described in Hristova and Wimley (2023) [50].

In order to assess viral compositional differences between methods, counts of reads assigned to each viral family were normalized by dividing by the total number of viral reads in each sample to get the relative abundances. The abundances were then transformed using the centered-log-ratio (CLR) transformation to account for the compositional nature of the data and allow for comparison between groups.

## 3. RESULTS

We used a baseline feces virome protocol (Protocol A; Figure 1) as the reference for improvement and tested several modifications at key methodological steps. The relative performance of the different protocols was evaluated using multiple parameters, including taxonomic and functional read assignments, contig characteristics and quality, viral diversity metrics, and genome type and structure classification of the viral families identified.

### 3.1 Overview of the outcome of the protocols tested

Pair-end read counts (or single reads counts) before and after QC and filtering, along with read assignments to human, viral and bacterial taxa—as well as unassigned reads—are summarized in Tables S1 and S2 for all protocols. Protocols without viral enrichment (A.0.1 and A.0.2) showed the highest proportions of assigned reads (80.96-96.85%; Mann-Whitney U-test p<0.05 for A.0.1), though more than 95% were bacterial reads and only a minor fraction corresponded to viruses (≤0.002%) (Figure 2A). In contrast, Protocol A, which includes viral enrichment, yielded 20.3 times more viral reads than A.0.1 and 14.7 times more than A.0.2. This higher viral recovery and lower bacterial contamination resulted in an improved virus-to-bacteria ratio for Protocol A (Mann-Whitney U-test p<0.05 for A.0.1) (Figure 2B).

**Figure 2.**
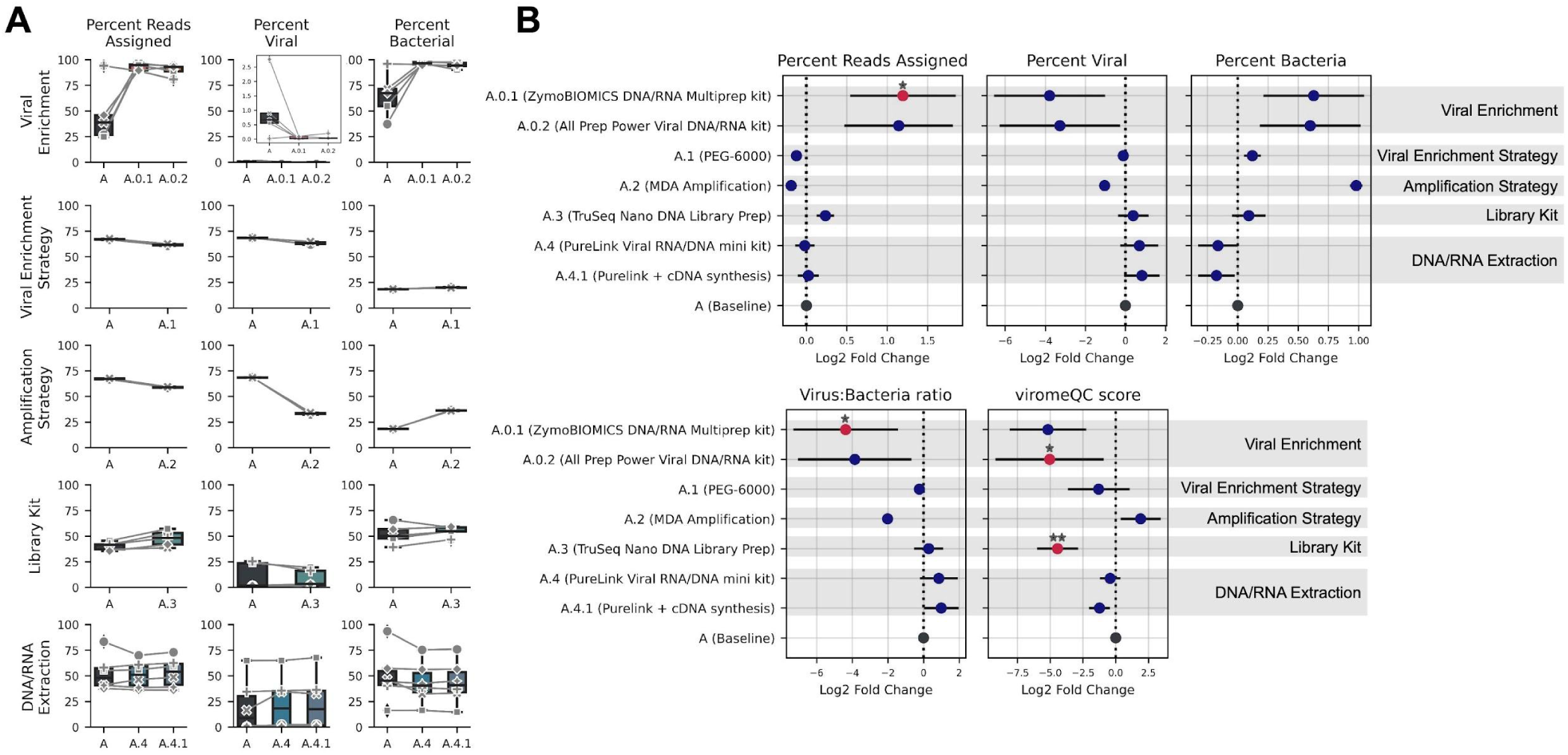
Overview of the outcomes of the tested protocols based on SeqScreen assignments. A) Boxplots showing the percentage of reads assigned to any taxa, viruses and bacteria for each protocol. The central line represents the median, box limits indicate Q1 and Q3, and whiskers extend to the lowest and highest non-outlier values (Q1-1.5 IQR and Q3+1.5 IQR). Individual sample data points are also displayed. B) Mean and 95% confidence interval (CI) of the log2 fold-change in read assignments (total reads, viral reads, bacterial reads), the virus-to-bacteria ratio, and the viromeQC score. Each modified protocol was compared to Protocol A within its respective set of processed samples. Significant differences are labelled with a star (p<=0.05, |fold change| > 1).

Ultracentrifugation (A) outperformed the PEG-based purification (A.1), resulting in a higher proportion of viral reads and reduced bacterial contamination (Figure 2). Despite the inclusion of a viral enrichment step, the inherently low viral nucleic acid yields make amplification a necessary step before the preparation of the sequencing libraries. The choice of the amplification strategy had a significant impact. Using MDA (A.2) instead of PCR-SISPA (A) halved the median proportion of viral reads (MDA=33.32% vs PCR-SISPA=68.45%) and doubled the median percentage of bacterial reads (MDA=36.30% vs PCR-SISPA=18.42%). The median virus-to-bacteria ratio for PCR-SISPA and MDA was 3.7 and 0.92, respectively, representing a ∼4-fold increase using PCR amplification (Figure 2). Interestingly, despite its lower viral recovery, MDA resulted in the highest viromeQC scores, suggesting a trade-off between viral enrichment and sequencing quality. The alignment rate for MDA and PCR-SISPA protocols were 0.0062 and 0.0047 for SSU rRNA, 0.0048 and 0.026 for LSU rRNA, and 0.00043 and 0.0021 for the bacterial markers, respectively.

Library preparation can influence sequencing sensitivity and virome diversity capture. We compared two standard methods for library generation: a ligation-based kit (A.3; TruSeq Nano DNA) and a tagmentation-based kit (A; DNA Prep (M) Tagmentation). Compared to protocol A, TruSeq Nano showed slightly higher median reads assigned to any taxa, including viruses and bacteria, resulting in marginally increased virus-to-bacteria ratios (Figure 2).

The nucleic acid extraction method is critical for maximizing viral RNA and DNA recovery. The QIAamp Viral RNA Mini kit (A) is widely used in virome studies (reviewed in Bassi et al. 2022 [51]). Since this kit is primarily designed for viral RNA extraction with DNA co-purification, we tested an alternative kit specifically optimized for both viral DNA and RNA extraction, the PureLink Viral RNA/DNA Mini kit (A.4). Using the PureLink kit resulted in a median two-fold increase in viral reads compared to the QIAamp kit, along with a minor reduction in bacterial contamination and 2.5-fold higher virus-to-bacteria ratios (Figure 2). We also tested the impact of double-stranded cDNA synthesis after retrotranscription (A.4.1). The viral read percentage and other metrics remained comparable between Protocol A.4 and A.4.1, suggesting that this additional step does not improve virome recovery.

### 3.2 Viral assembly quality

We also evaluated the performance of the protocols based on the characteristics and quality of viral assemblies. Samples processed with protocols A.0.1 and A.0.2 (without viral enrichment) generated more and longer contigs compared to protocol A (which includes viral enrichment) (Figure 3). However, CheckV quality estimates classified all contigs from A.0.1 and A.0.2 as either non-viral or of low quality, whereas protocol A produced contigs of medium and high quality, including complete genomes.

**Figure 3.**
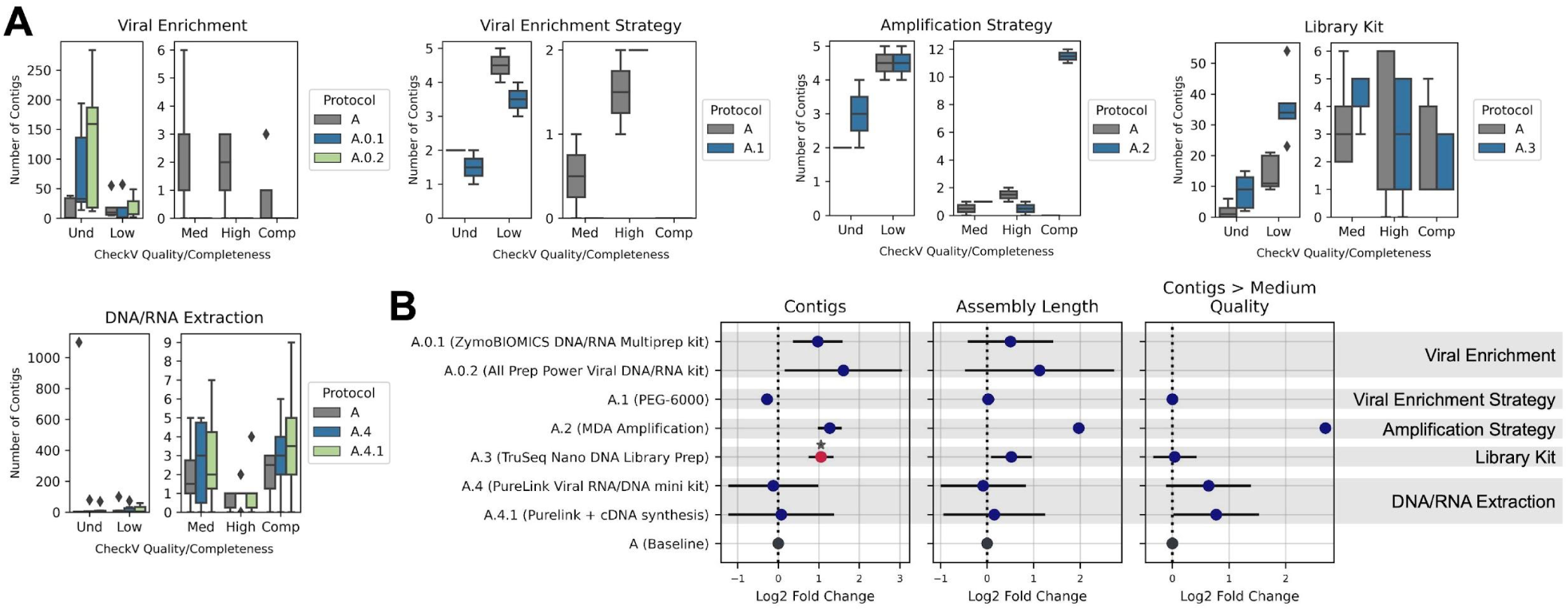
Quality of viral assemblies across the protocols tested. A) Boxplots showing the number of contigs >1000 bp assembled using MEGAHIT, categorized by CheckV quality and completeness scores: *Und*: undetermined quality (no completeness estimate available), *Low* quality: <50% completeness, medium (*Med*) quality: 50-90% completeness, *High* quality: ≥90% completeness, and complete genomes (*Comp*). The central line represents the median, box limits indicate the Q1 and Q3 quartiles, and whiskers extend to Q1-1.5 IQR and Q3+1.5 IQR, marking the lowest and highest non-outlier values. B) Mean and 95% CI of the log2 fold-change in the number of contigs >1000 bp, assembly length (total nucleotides in those contigs) and total number of contigs classified as medium, high-quality, or complete. Each modified protocol was compared to Protocol A within its respective set of processed samples. Significant differences are labelled with a star (p<=0.05, |fold change| > 1). Protocols A.0.1 and A.0.2 do not have contigs at medium or greater quality.

Compared to the ultracentrifugation enrichment (Protocol A), PEG-based precipitation (Protocol A.1) resulted in fewer, shorter and lower-quality contigs, with a median length of 931.5 pb for A.1 and 1440 pb for A. The amplification strategy also had a notable impact on viral assembly metrics. MDA-amplified samples (Protocol A.2) yielded more and longer contigs than PCR-SISPA-amplified samples (Protocol A). Notably, protocol A.2 produced a higher number of contigs above 1000 bp and an increased proportion of high-quality assemblies, particularly of complete genomes (Figure 3). However, CheckV predictions on closed genomes indicated that all 11 and 12 predicted complete genomes from the two samples processed with protocol A.2 contained direct terminal repeats (DTR), suggesting that these corresponded to circular viral genomes [47].

Several differences were also observed in the library preparation strategies. The ligation-based approach (Protocol A.3; TruSeq Nano) generated longer contigs compared to the tagmentation-based approach (Protocol A; DNA Prep (M) Tagmentation) (Mann-Whitney U-test p<0.05). While the total number of medium-quality contigs was slightly higher for TruSeq Nano libraries, DNA Prep libraries produced more high-quality contigs and complete genomes. Similar to protocol A.2, all complete genomes obtained with protocol A.3 contained DTRs, indicative of circular viral genomes.

Lastly, while the nucleic acid extraction protocols (A, A.4 and A.4.1) resulted in similar contig numbers and lengths, their completeness estimates varied. Protocols A.4 (PureLink kit) and A.4.1 (PureLink + ds-cDNA synthesis) produced a higher proportion of medium-quality contigs and complete genomes compared to protocol A (QIAamp kit).

### 3.3 Richness and alpha diversity metrics

Richness and diversity are fundamental metrics in virome studies, as they reflect a protocol’s ability to capture virome complexity. The inclusion of a viral enrichment step (Protocol A) significantly increased taxonomic richness (p<1e-7 compared to protocols A.0.1 and A.0.2, at family level), leading to the detection of a greater number of unique viral families and species compared to direct nucleic acid extraction without viral enrichment (Figure 4A). This suggests that viral enrichment enhances viral genome recovery, even at thresholds under 200 reads (Figure S1). A similar trend was observed for diversity, with protocol A showing a more diverse and evenly distributed virome as indicated by both Shannon and Simpson indices (p<1e-10 at family level) when compared to protocols A.0.1 and A.0.2 (Figure 4B and 4C).

**Figure 4.**
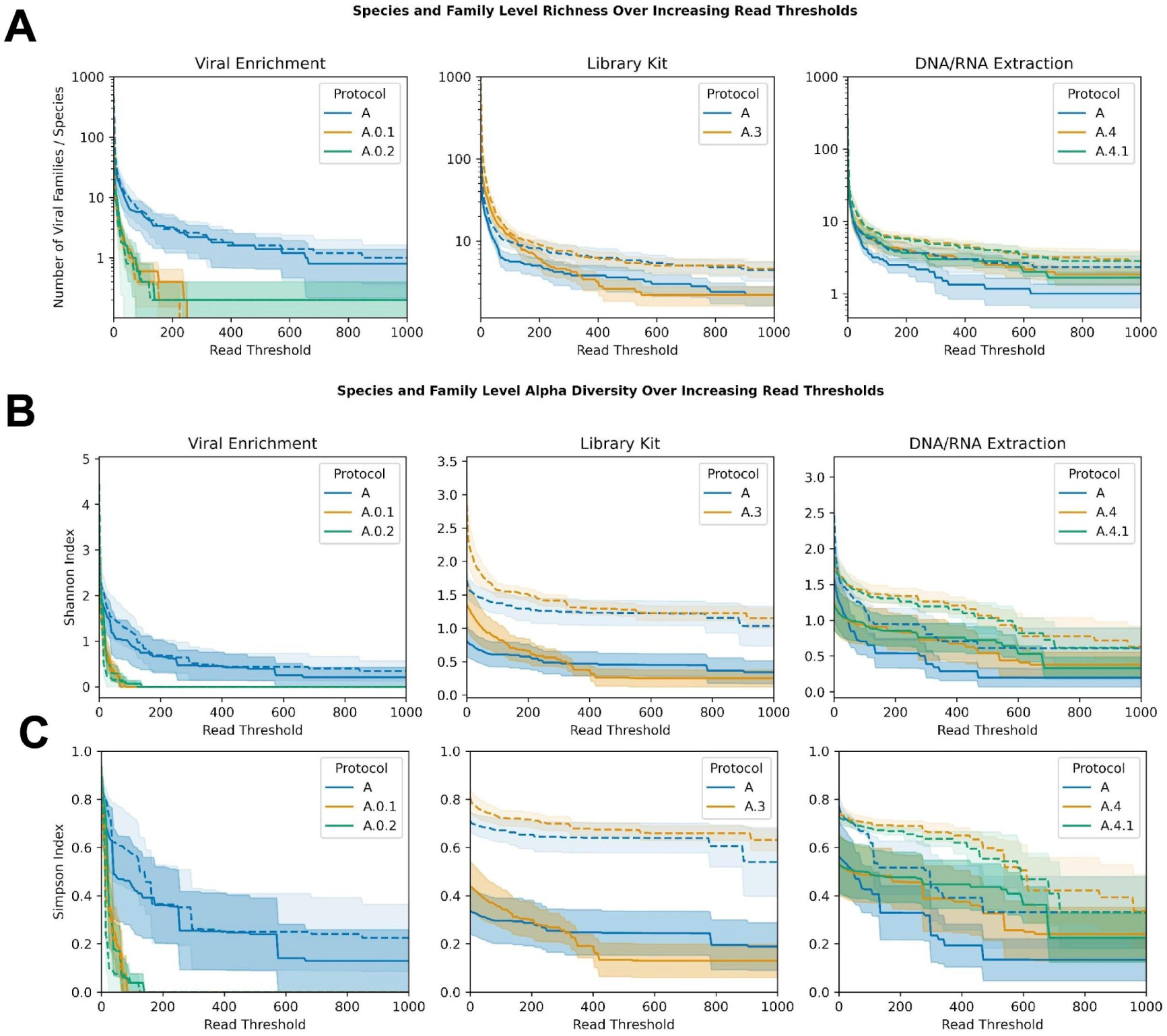
Richness and diversity of viral taxonomic assignments across protocols. Species-level metrics are shown with dashed lines, while continuous lines represent family-level metrics. Each metric is calculated at increasing read thresholds, disregarding any taxa below the required number of reads. Cutoffs from 0-1000 with step sizes of 5 were calculated. A) Species-and family-level richness. B) Species-and family-level Shannon index. C) Species-and family-level Simpson index. Protocols A.1 and A.2 were excluded from the analysis due to the small number of samples (n=2).

Library preparation had a measurable impact on viral diversity (Figure 4 and Figure S1). The tagmentation-based approach (Protocol A) showed slightly higher richness and diversity than the ligation-based preparation (Protocol A.3) at read thresholds above 350, whereas the latter outperformed at thresholds below 200. The Shannon index values suggest that protocol A captured greater diversity and evenness at the family level above 350 read thresholds, while at species level, diversity remained comparable between the two protocols (Figure 4B). A similar trend was observed for the Simpson index, with protocol A exhibiting lower dominance and greater evenness at the family level. However, at the species level, the Simpson index values were slightly higher for protocol A.3, suggesting a marginally more balanced distribution of species in this library preparation method.

The choice of the nucleic acid extraction kit also influenced viral richness and diversity. The PureLink kit (Protocols A.4 and A.4.1) showed higher richness of viral taxa (p<1e-5 for protocol A.4.1 at family level) compared to the QIAamp kit (Protocol A) (Figure 4A). Higher diversity metrics were also found above 50 read thresholds (Figure 4B and 4C, and Figure S1), resulting in significant differences for Shannon and Simpson indices (p < 1e-4 at family level) regarding protocol A.4.1. No major differences were found between protocols A.4 and A.4.1, indicating that the addition of a second-strand synthesis step did not substantially impact richness or diversity.

Richness and diversity metrics were not assessed for protocols A.1 and A.2 (PEG-600 precipitation and MDA amplification, respectively) due to the small number of samples processed (n=2).

### 3.4 Genome composition and structure of the assigned viruses

The diversity and complexity of viral particles raise the question of whether specific methodological choices selectively recover viruses with particular genomic and structural characteristics, potentially influencing the virome representation. To address this, we examined family-level taxonomic assignments and assessed key viral traits, including genome type and strandness (DNA or RNA, single-or double-stranded), genome structure (linear or circular), genome size, viral particle size, and the presence or absence of an outer envelope.

Across all protocols, in most of the samples processed, we observed a high abundance of viruses with circular genomes (median 76.48%), ssDNA (median 66.87%) (Figure S2), smaller than 15 kb (median 89.28%), non-enveloped (median 93.08%), and ≤50 nm in size (median 66.89%). This trend was largely driven by the *Microviridae* family, which possesses these characteristics and accounted for nearly 50% of the viral reads on average, with values ranging from 0 to 99.75% across samples. To assess how protocols influence viral capture beyond this family, we reanalyzed the data after excluding the *Microviridae* reads (Figure 5 and Figure S3).

**Figure 5.**
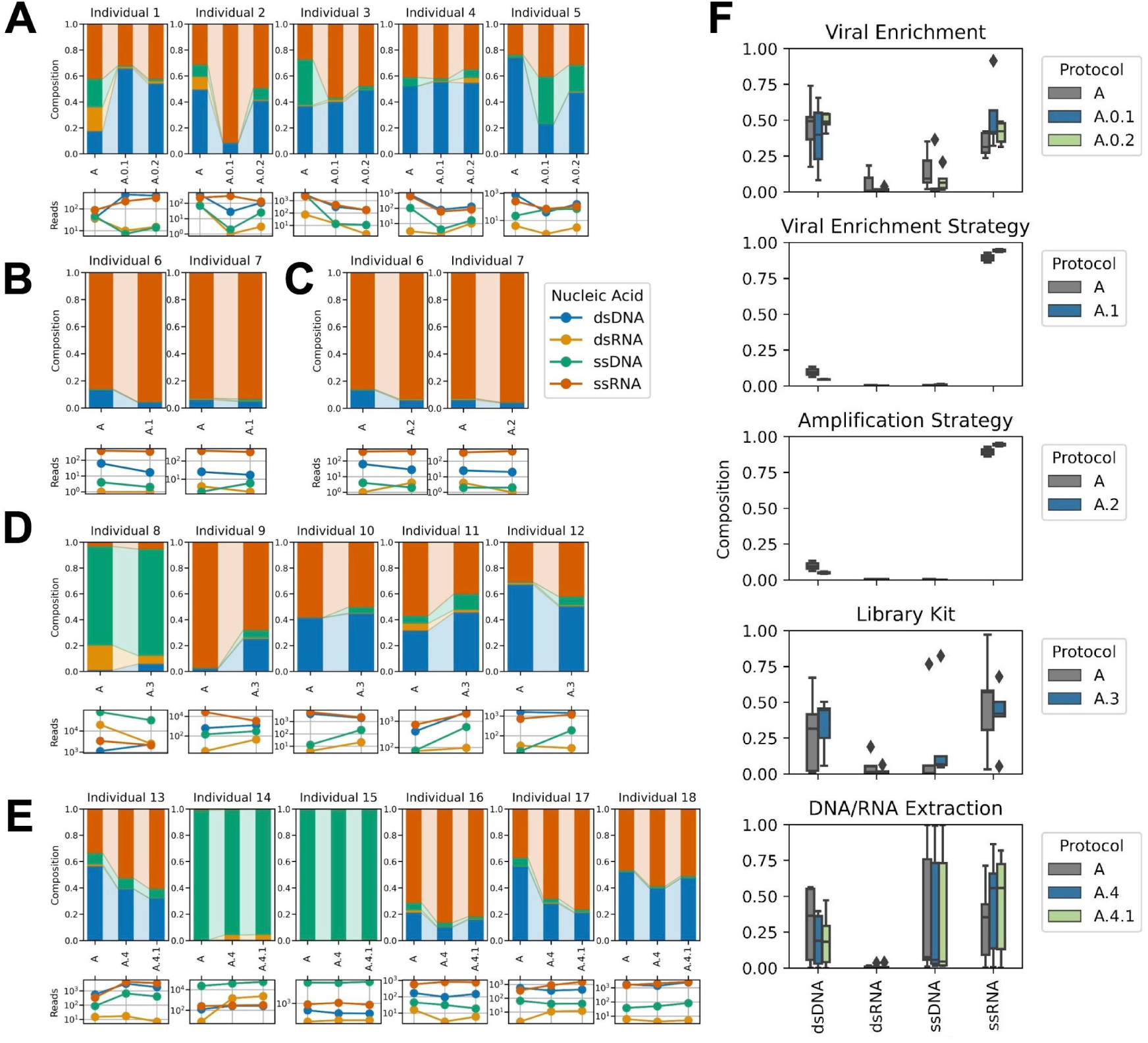
Diversity and composition of viruses by genome type for the protocols tested after excluding the *Microviridae* reads. For A-E, the top figures represent the genome composition of individual samples, while the bottom ones show the reads of genome types (dsDNA, ssDNA, dsRNA and ssRNA). A) Viral enrichment, where protocol A corresponds to viral precipitation by ultracentrifugation and protocols A.0.1 and A.0.2 lack viral enrichment step (nucleic acids were directly extracted using ZymoBIOMICS DNA/RNA Mini kit or AllPrep Power Viral DNA/RNA kit, respectively). B) Viral enrichment strategy, where protocol A corresponds to ultracentrifugation and protocol A.1 to PEG-600-based precipitation. C) Amplification strategy, where protocol A corresponds to PCR-SISPA and protocol A.2 to MDA. D) Library preparation kit, where protocol A corresponds to the DNA Prep (M) Tagmentation kit and protocol A.3 to the TruSeq Nano DNA Library Prep kit. E) DNA/RNA extraction, where protocol A corresponds to the QIAamp Viral RNA Mini kit, protocol A.4 to the PureLink Viral RNA/DNA Mini kit and protocol A.4.1 to PureLink Viral RNA/DNA Mini kit followed by the cDNA second-strand synthesis. F) Boxplots showing the genome composition for the protocols. The central line represents the median, box limits indicate the Q1 and Q3 quartiles, and whiskers extend to Q1-1.5 IQR and Q3+1.5 IQR, marking the lowest and highest non-outlier values.

The samples processed with protocol A.0.1 showed a lower median proportion of dsDNA viruses (39.95%) compared to protocol A.0.2 (49.16%) and protocol A (49.53%) (Figure 5A and 5F). However, protocol A captured a higher proportion of ssDNA and dsRNA viruses, while protocol A.0.1 recovered the largest proportion of ssRNA viruses, predominantly ssRNA-RT (Figure S3). All three protocols primarily recovered linear genomes <15 kb, although protocol A assigned >30% of reads to families with genomes >50 kb. Regardless of the protocol, most viral particles were non-enveloped and ranged from 50 to 200 nm in size. More than 20% of families corresponded to phages in protocol A (Figure S4).

The choice of the viral enrichment method, PEG-based precipitation (A.1) or ultracentrifugation (A), resulted in similar trends across viral genome characteristics evaluated in the two samples processed, with ssRNA genomes representing ∼90% of the recovered families (Figure 5B and 5F), mainly ssRNA(+) (Figure S3). In both cases, the most abundant viruses had linear genomes <15 kb, were enveloped, and measured <50 nm. Phages accounted for 1.38% (A.1) and 7.25% (A) of the recovered families (Figure S4).

Regardless of the amplification strategy—MDA (A.2) or PCR-SISPA (A)—the majority of recovered genomes were ssRNA, with less than 10% corresponding to dsDNA (Figure 5C). Linear genomes <15 kb and under 50 nm were the most abundant in both cases, but MDA primarily captured non-enveloped viruses (median 78.69%), whereas PCR-SISPA predominantly recovered enveloped viral particles (median 86.75%). The median proportion of phages was 2.50% for protocol A.2 and 7.25% for protocol A (Figure S4).

The viral family composition remained consistent between ligation-based (A.3) and tagmentation-based (A) libraries (Figure 5D). For both protocols, the proportions of dsDNA, ssDNA, dsRNA, and ssRNA averaged ∼31%, ∼19%, ∼3%, and ∼45%, respectively (Figure 5F). Over 60% of the genomes were linear and smaller than 15 kb, viruses were non-enveloped and primarily <200 nm in size, and phages accounted for less than 10% (Figure S4).

We observed differences in genome composition between nucleic acid extraction kits (A vs. A.4), but genome type proportions were comparable between the inclusion or omission of a second-strand cDNA synthesis step (A.4.1 vs. A.4) (Figure 5E and Figure S3). While protocol A (QIAamp kit) captured dsDNA, ssDNA, and ssRNA genomes in similar proportions, protocols A.4 and A.4.1 (PureLink kit) showed a higher abundance of ssRNA viruses (∼45%), followed by ssDNA (∼35%) and dsDNA (∼20%) (Figure 5F). In all cases, recovered genomes were predominantly linear and <15 kb, corresponded to non-enveloped viruses, and phages accounted for less than 10% of the viral reads. In terms of viral particle size, more than 50% of the viruses recovered with A.4 and A.4.1 were >200 nm, a proportion notably higher than in protocol A (Figure S3). Differences are also evidenced in the 25 most frequent viral families detected in protocols A, A.4 and A.4.1 (Figure 6).

**Figure 6.**
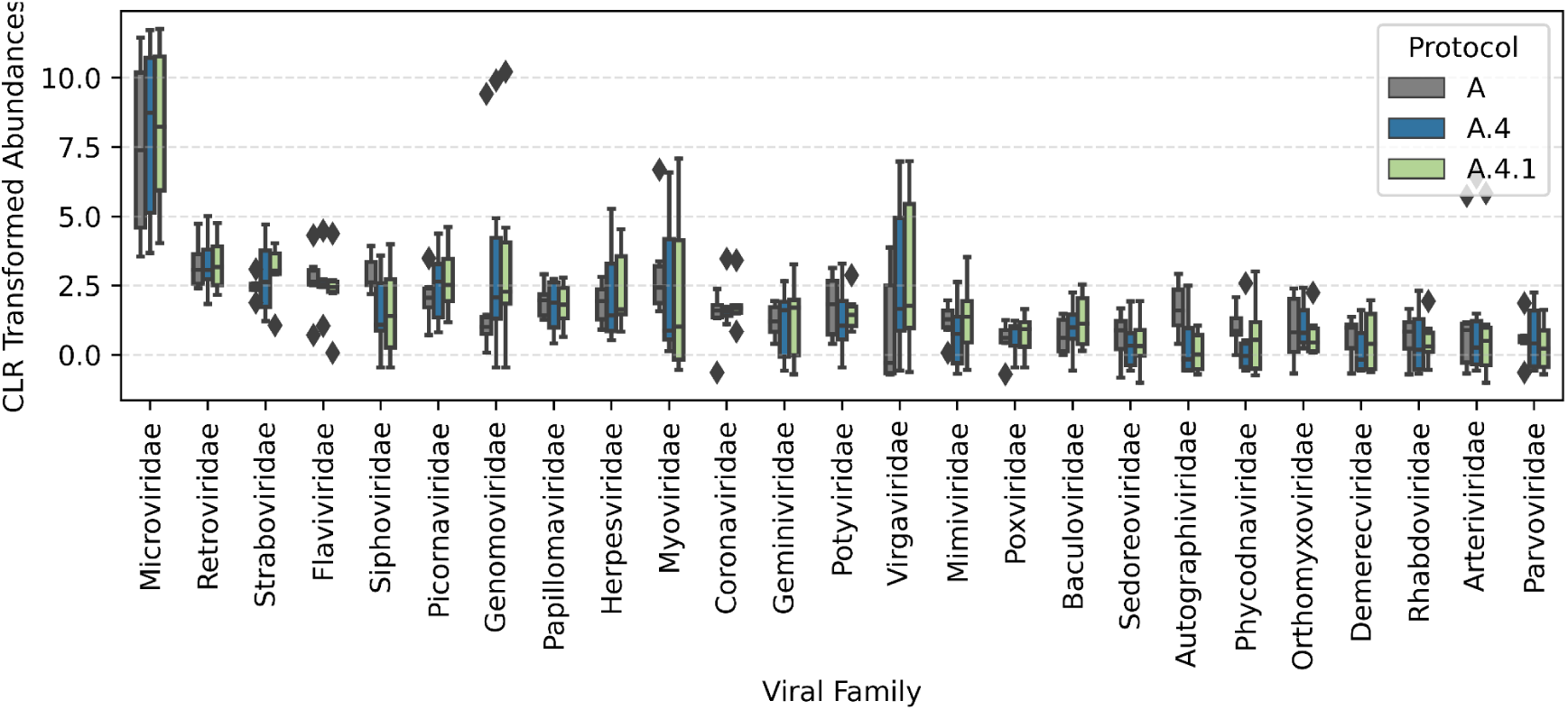
Most frequent viral families for the nucleic acid extraction protocols tested. Viral counts at the family level were compositionally transformed before applying the centered-log-ratio (CLR) transform to the data. The 25 families with the highest median CLR values are displayed. Protocol A corresponds to the QIAamp Viral RNA Mini kit, protocol A.4 to the PureLink Viral RNA/DNA Mini kit and protocol A.4.1 to the PureLink Viral RNA/DNA Mini kit followed by the cDNA second-strand synthesis.

## 4. DISCUSSION

The study of the human virome has gained increasing attention in recent years due to its crucial role in human health. The gut virome, in particular, is a highly diverse and dynamic ecosystem, predominantly composed of bacteriophages that influence bacterial populations, their metabolism, and host immunity [11–13,15]. Viromes derived from fecal samples are commonly used as a proxy for the gut virome and can provide valuable insights into gut-associated viral communities, but methodological challenges remain [52]. Viruses lack universal genetic markers, making taxonomic classification difficult, and a large proportion of viral sequences remain uncharacterized, contributing to the so-called “viral dark matter” [22–24]. Additionally, virome analysis is complicated by low viral biomass in clinical samples, contamination from host and bacterial nucleic acids, and biases introduced by sample processing methods. Therefore, optimizing virome sequencing protocols is essential to improve viral recovery, reduce contaminants, and achieve a more accurate representation of viral communities, contributing to reproducible results that can lead to more reliable biological conclusions.

In this study, we evaluated the impact of different methodological modifications in a fecal virome metagenomics protocol on multiple parameters, including taxonomic and functional read assignments, contig characteristics and quality, viral diversity metrics, and genome type and structure classifications.

Protocol A, which was used as the reference for testing several modifications, was designed by combining and adapting certain steps from protocols and technical recommendations [29–31,34,53]. The first step of protocol A is homogenization and lysis through bead-beating using 0.1 and 0.5 mm beads. Since gut viruses exist in multiple states —free virions, intracellular replicating viruses, attached to host or microbial cells, and as dormant proviruses or episomes [14]— we implemented bead-beating to enhance viral recovery by lysing bacterial and host cells, detaching viruses attached from biofilms, and reducing contaminating host DNA. The effect of bead size was previously evaluated using a virus mock community, demonstrating a notable decrease in certain viral particles when 2.8 mm beads were used instead of 0.1 mm beads [30].

Eukaryotic and most microbial genomes are significantly larger than viral genomes, often dominating the total nucleic acid yield in a sample, making viral enrichment and precipitation crucial in virome studies [30,51,54]. Several approaches, including low-speed centrifugation, filtration, nuclease digestion, CsCl density gradient centrifugation, ultracentrifugation, tangential flow filtration and PEG, have been used in different combinations [29–31,34,53,55–58]. The choice of purification methods can impact viral recovery due to the vast diversity of viral structures, sizes, genome composition, and resistance to chemical and mechanical stressors. Following low-speed centrifugation, we used 0.45 μm filters instead of 0.2 μm filters used in previous studies [55,59,60]. The microscopic examination of filtered feces through 0.2 μm pores showed a close to 50% reduction of viral particles while no bacterial cells were observed, highlighting the trade-off between bacterial removal and viral particle retention [61]. Given that certain Myoviridae and Siphoviridae phages and giant eukaryotic viruses exceed 0.2 μm in size [29,62], using 0.45 μm filters ensures the retention of larger viruses while still removing bacterial and host cells.

The recent development of kits for microbial metagenomics prompted us to explore whether we could avoid the time-consuming viral enrichment steps by directly extracting microbial nucleic acids. Despite incorporating the modifications recommended from both providers for viruses, the ZymoBIOMICS DNA/RNA Miniprep kit (Protocol A.0.1) and the AllPrep PowerViral DNA/RNA kit (Protocol A.0.2) performed poorly, yielding ∼15-20 times fewer viral reads compared to protocol A, significantly reducing viral diversity. Although protocols A.0.1 and A.0.2 produced larger contigs, the CheckV analysis indicated they were of low quality or possibly non-viral in origin, confirming that metagenomic DNA/RNA extraction kits designed for bacteria are unsuitable for virome studies.

Since viral enrichment seems compulsory, we tested PEG precipitation (Protocol A.1) as an alternative to ultracentrifugation (Protocol A), which requires high-quality equipment that may not be available in smaller laboratories. PEG has been used before [31,57,58,61] due to its ability to sequester water molecules, forcing the aggregation of viruses, and being scalable to large volumes of sample, allowing high-throughput. On the other hand, ultracentrifugation with pellet resuspension or CsCl density gradient remains a cornerstone in virology for purifying viruses from viral lysates from cell cultures [63], environmental [64,65], wastewater [66] and clinical samples [67]. CsCl density gradient is labor-intensive and poorly reproducible as it requires precise gradient preparation and careful fraction collection to purify viruses within specific density ranges [29], introducing variability between experiments. Therefore, we decided to use ultracentrifugation with pellet resuspension. We found minor differences between PEG and ultracentrifugation, with median virus-to-bacteria ratios of 3.16 and 3.72, respectively, recovering viruses that had similar genome characteristics and structure. However, PEG-based precipitation showed fewer total contigs and fewer contigs of at least medium quality compared to ultracentrifugation. Moreover, we had difficulties to pellet the PEG-virus complex since the supernatant had viscous remnants and we were unable to recover all the precipitated viruses, making it poorly reproducible. These difficulties were also found by Kleiner et al. (2015) [29], who suggested that chloroform could help remove residual PEG. However, chloroform can disrupt the phospholipid membranes of chloroform-sensitive enveloped viruses, such as dsRNA phages from the family Cystoviridae, potentially introducing bias into the study [28,31,55].

Free host/bacterial DNA can make up over 90% of reads in shotgun metagenomics, so nuclease digestion is a critical step in virome protocols. During this step, free host and bacterial nucleic acids (including free prophage DNA) are removed, preserving only intact viral genomes inside capsids. Since we are interested in both DNA and RNA viruses, our digestion strategy using Turbo DNAse, RNAse A, RNAse T1, and benzonase effectively reduced non-viral contamination, increasing the proportion of viral reads, consistent with previous studies [30,53].

Another relevant step in virome studies is the amplification of viral nucleic acids, often necessary due to the typical low yields of viral DNA and RNA. Amplification strategies can include a reverse transcription step to produce cDNA for sequencing of viral RNA genomes. Several methods for nucleic acid amplification, together with their potential biases, have been evaluated [68], including whole genome (WGA) or whole transcriptome amplification (WTA) [30,62,69–72], random priming PCR or single primer PCR (SISPA) [34,54,62,72–74], Rolling Circle Amplification (RCA) [75–77], and virus discovery based on cDNA-AFLP (VIDISCA) [78,79], among others.

We decided to focus on WGA using MDA with random hexamers (Protocol A.2) and PCR-SISPA (Protocol A), as these methods are widely used. Our PCR-SISPA approach relies on the D2_8N pseudo-degenerate oligonucleotide, containing 8 random nucleotides at the 3′ end for random priming and 20 nucleotides of defined sequence at the 5′ end, for retrotranscription of RNA to cDNA, and the D2 oligonucleotide for PCR-SISPA amplification using a high-fidelity DNA polymerase (Q5) that has a robust performance. Instead, MDA is not PCR-based, it amplifies DNA under isothermal conditions and relies on the high-fidelity and strong strand-displacement capacity of the phi29 polymerase to amplify ultralow amounts of DNA template [80]. We used the GenomiPhi v2 DNA Amplification kit, as it is one of the most used kits in virome studies [31,53,54].

With PCR-SISPA we achieved a median two-fold increase in viral reads, together with decreased bacteria reads, resulting in a four-fold superior virus-to-bacteria ratio for PCR. Despite these results, MDA samples showed the best ViromeQC scores. Sun et al. (2022) [71] evaluated bacterial contamination and found that MDA libraries had the lowest alignment rates to SSU and LSU rRNA but higher bacterial marker alignment rates compared to SISPA. This suggests that while ViromeQC detects contamination, interpreting its scores may depend on the amplification method employed.

Regarding viral assembly, MDA showed an increased number and length of contigs, with a significantly higher proportion of complete genomes. However, CheckV identified repeated sequences at both ends of the complete contigs, corresponding to DTRs [47]. These terminal repeats are consistent with circular viral DNA molecules, resulting from the assembly of short reads [81]. Better assembly metrics were reported by Parras-Moltó and colleagues (2018) [62] for MDA compared to SISPA, but the latter showed a more balanced genome representation with high-coverage peaks in certain genomic regions. Haagmans et al. (2025) [72] also found higher assembly quality and longer contigs when using the WTA2 kit, whereas SISPA resulted in more fragmented assemblies. Additionally, the comparison between MDA and non-MDA datasets included in the Gut Virome Database (GVD) did not reveal differences in viral recovery or number of contigs assembled, though studies relying on MDA (96%) were significantly enriched in eukaryotic, ssDNA viruses [82].

The bias on the preferential amplification of small circular ssDNA genomes is supported by the fact that the phi29 polymerase and its strand-displacement activity allow the amplification of circular genomes more efficiently than linear or larger dsDNA genomes [83,84]. In the case of SISPA, an annealing bias has also been reported, related to the defined part of the oligonucleotide, resulting in the uneven distribution of sequence reads across the genomes and affecting the sensitivity of detection for low-abundance viruses [85]. To reduce amplification biases, several studies based on MDA (including ours), performed pooling of three to five replicates of the amplification reaction for each sample [31,54,86]. Similarly, for SISPA the pooling of amplified products obtained with different primers has been suggested, providing more uniform coverage patterns [62].

A consequence of the biased amplification of circular ssDNA genomes has been linked to the overrepresentation of certain viral families, such as *Microviridae* [28,62]. We found a close to 50% abundance of reads assigned to *Microviridae* in both samples, and more than 90% of the assigned families had circular genomes, regardless of the amplification protocol used. Therefore, it remains unclear whether the predominance of circular genomes is real or a result of methodological bias.

Library preparation methods for virome studies also lack standardization and have not been evaluated thoroughly despite their influence on the detection and characterization of viral communities. Illumina short-read sequencing is mostly used, however, long-read sequencing (Oxford Nanopore Technologies (ONT) or PacBio) is also starting to be implemented in viral metagenomics [87,88]. A recent study comparing the three technologies reported that Illumina performed best at genome recovery and had the lowest error rates, but the combination of Illumina and ONT (hybrid assembly approach) improved the assembly of low-abundant genomes while reducing ONT-related errors [89]. An inconvenience for ONT-only sequencing is the need for lengthy amplification products as input for the library, relying on MDA [54,89].

In our study, we focused on Illumina sequencing and evaluated two library kits that include PCR and are recommended for shotgun metagenomic sequencing. While the DNA Prep (M) Tagmentation kit (Protocol A) relies on on-bead tagmentation chemistry which simultaneously fragments DNA and adds adapters, requires 1-500 ng DNA for small genomes and has a target insert size of ∼350 bp, the TruSeq Nano DNA Library Prep (Protocol A.3) implements mechanical DNA fragmentation and adapter ligation, the input is 100 ng of genomic DNA and the insert size is 350 bp or 550 bp. However, we adapted the TruSeq library preparation protocol and skipped fragmentation since our input was PCR-SISPA products (200-1000 bp), not genomic DNA. Both kits generated functional libraries, and subtle differences were found for some of the parameters evaluated. Viral reads and virus-to-bacteria ratios were moderately increased for TruSeq libraries, as well as medium-quality contigs. However, we found that all the complete genomes from this library presumably corresponded to circular viral genomes, possibly reflecting a methodological bias inherent to library preparation, which should be further evaluated. On the other hand, the tagmentation-based kit resulted in more contigs classified as complete genomes, and higher richness and diversity metrics at the family level, especially for read thresholds over 400. Genome composition was comparable among kits, with *Microviridae* as the most abundant viral family in all the samples, independently of the library kit. Excluding this ssDNA family, ssRNA viruses were highly abundant, followed by dsDNA, other non-*Microviridae* ssDNA viruses, and only a minor proportion corresponding to dsRNA viral genomes. The genome structure of the viruses assigned was also similar between library kits, with linear genomes <15 kb corresponding to <200 nm non-enveloped viruses prevailing.

To our knowledge, no study has compared Illumina library kits for the study of the human virome. However, Sato et al. (2019) [90] compared the sequencing bias introduced by different Illumina library preparation kits for bacteria metagenomics. They reported that Nextera DNA Flex (former name of the DNA Prep (M) Tagmentation kit we used) exhibited less severe GC bias than Nextera XT (widely used in virome studies [31,34,60,74,89]), but still showed some GC content-associated sequencing bias, particularly in low-GC regions. Concerning TruSeq Nano, they found more uniform sequencing coverage, but some sequencing bias was introduced in extreme GC-content regions. They found Nextera DNA Flex faster and more efficient for low-input DNA samples, but it can introduce sequencing bias that affects relative abundance estimates in metagenomic datasets. In our study, the five samples sequenced with both kits showed similar average %GC content, ranging from 41-45% for DNA Prep and 46-50% for TruSeq. These values align with the two main GC-content peaks (∼40% and ∼50%) reported in the metagenomic catalog of the human gut virome [91], where the majority of viral sequences are concentrated.

In all the protocols evaluated we extracted viral RNA and co-purified DNA using the QIAamp Viral RNA Mini kit (Protocol A), which according to the systematic review by Bassi et al. (2022) [51] is the most employed commercial kit for viral DNA and RNA extraction. As an alternative, we tested the PureLink Viral RNA/DNA Mini Kit (protocols A.4 and A.4.1). This kit outperformed the Qiagen kit in all the parameters evaluated. Reads assigned to viruses increased two-fold while bacteria reads decreased 0.9-fold, reaching a 2.5-fold increase in the virus-to-bacteria ratio with the Purelink kit. Also, the number of contigs classified as medium-, high-quality or complete was 1.7-fold superior for the Purelink kit, as well as higher richness and diversity metrics at family and species level. *Microviridae* was not predominantly the most abundant family in the 6 samples processed for this comparison, but excluding this family, we found that ssRNA viruses were more abundant for Purelink samples while ssDNA viruses were in higher proportion with the Qiagen kit. Genome structure was comparable with the two extraction kits, with a higher prevalence of linear genomes smaller than 15 kb, mainly non-enveloped and larger than 200 nm. Since the addition of a double-stranded cDNA synthesis step (protocol A.4.1) rendered merely similar results as protocol A.4, we decided to dispense this step in our final, optimized protocol.

Lewandowska and colleagues (2017) [92] compared the QIAamp Viral RNA Mini Kit, PureLink Viral RNA/DNA Mini Kit, and the automated NucliSENS EasyMAG system, and reported EasyMAG extraction was most efficient for RNA viruses. Regarding DNA viruses, concentrations were similar between the automated system and PureLink. They also highlighted that the Qiagen extraction kit led to the lowest recovery of viral genomes for all tested viruses. On the contrary, Klenner et al. (2017) [93] compared eight extraction kits, including the ones we tested, and found greater DNA yields for the Qiagen kit.

A relevant problem in metagenomic studies are external contaminants, which seem to come from nucleic acid extraction kits and also from WGA kits (DNA and RNA), contributing to the “kitome”, which refers to signals (reads) derived from reagent contaminants and kits, that are largely indistinguishable from microbiome signals and mostly have their unique profile specific to particular reagents and kits [94]. While bacteria are the most common contaminants, viruses have also been found in extraction kits [95]. The inclusion of no template controls (NTC) is of great importance to detect sporadic or reagent-related contaminants in the sequencing data. In our study, we prepared three independent NTC, composed of 250 μL of HBSS, that were processed in parallel to the feces samples, using our optimized protocol. Sequencing reads were processed following the same bioinformatic pipeline as the samples. SeqScreen assigned 83.33%, 56.21% and 83.32% of the reads in the three NTC, with more than 70% assigned to bacteria and less than 1% to virus. The number of viral families assigned with more than 100 reads were as follows: 7 families in NTC-1, 10 in NTC-2 and 1 in NTC-3. Picornaviridae was the only family that was common in the three NTC, with 145 reads in NTC-1, 285 reads in NTC-2 and 199 reads in NTC-3. The inclusion of more NTC could help determine whether contaminants are sporadic or related to a specific reagent or kit. While Salter et al. (2014) [96] prepared an extensive list of potentially contaminating bacteria genera related to sample processing (16S and bacteria metagenomic sequencing), no such list has been reported yet for virome metagenomics.

Several limitations should be acknowledged. First, we did not use the same fecal samples across all protocol modifications, which could introduce variability in the comparisons. Second, some methodological modifications, such as MDA amplification (Protocol A.2) and ligation-based library preparation (Protocol A.3), were tested on only two samples, limiting results. Additionally, our taxonomic and functional assignments were based on SeqScreen, which, while effective, may introduce biases in the classification of viral sequences. Furthermore, while our protocol enhances viral genome recovery and diversity representation, further validation in larger, independent cohorts is necessary to confirm its robustness and applicability.

Based on all of the above, our optimized protocol consists of bead-beating homogenization, viral enrichment (low-speed centrifugation, filtration with 0.45 μm pore size, ultracentrifugation, and nuclease digestion), RNA and DNA extraction using PureLink Viral RNA/DNA Mini Kit, RNA retrotranscription, PCR-SISPA amplification using pseudo-degenerate oligonucleotides, and sequencing library preparation using an on-bead tagmentation approach (Protocol A.4). This protocol significantly improved viral genome recovery, minimizing contamination from non-viral sequences, and enhanced the generation of high-quality contigs. Moreover, it resulted in increased viral richness and diversity, capturing DNA and RNA viruses, both double-and single-stranded, offering a more comprehensive and accurate representation of the gut virome.

## 5. CONCLUSION

Our optimized protocol for fecal virome analysis significantly improves viral recovery, minimizes host and bacterial contamination, and enhances viral diversity and genome completeness. Our findings highlight the importance of viral enrichment, the advantages of PureLink extraction, the effectiveness of PCR-SISPA over MDA, and the superior performance of tagmentation-based library preparation. These results contribute to advancing virome research by providing a robust and reproducible approach for studying the gut virome in human feces. Future studies should further validate these findings across larger cohorts and explore additional refinements to virome sequencing methodologies.

## Data Availability

FASTQ files generated for this study are available in the BioProject repository with the accession number PRJNA1250239. All code and scripts used for bioinformatic processing, taxonomic assignment, assembly, richness and diversity analysis, and statistical analyses are available at: https://github.com/rdoughty10/GutVirome.

## 6. ACKNOWLEDGEMENTS

This project received funding from *“Fundación Científica de la Asociación Española contra el Cáncer”* (PRYGN211425POSA), granted to D.P. R.D. is supported by a training fellowship from the Gulf Coast Consortia, on the NLM Training Program in Biomedical Informatics & Data Science (T15LM007093).

T.J.T is supported in part by the National Institutes of Health, National Institute of Allergy and Infectious Diseases (P01-AI152999, U19-AI144297) and the National Science Foundation (EF-2126387, IIS-2239114).

## 8. SUPPLEMENTARY MATERIAL

**Table S1:** Read counts before and after QC and filtering.

**Table S2:** Summary of taxonomic read assignment.

**Figure S1:** Richness and diversity of viral taxonomic assignments for read thresholds below 200. Species-level metrics are shown with dashed lines, while continuous lines represent family-level metrics. Each metric is calculated at increasing read thresholds, disregarding any taxa below the required number of reads. Cutoffs from 0-200 with step sizes of 5 were calculated. A) Species-and family-level richness. B) Species-and family-level Shannon index. C) Species-and family-level Simpson index. Protocols A.1 and A.2 were excluded from the analysis due to the small number of samples (n=2).

**Figure S2:** Composition of viruses by genome type for the protocols tested. For A-E, the top figures represent the genome composition of individual samples, while the bottom one show the reads of genome types (dsDNA, ssDNA, dsRNA and ssRNA). A) Viral enrichment, where protocol A corresponds to viral precipitation by ultracentrifugation and protocols A.0.1 and A.0.2 lack viral enrichment step (nucleic acids were directly extracted using ZymoBIOMICS DNA/RNA Mini kit or AllPrep Power Viral DNA/RNA kit, respectively). B) Viral enrichment strategy, where protocol A corresponds to ultracentrifugation and protocol A.1 to PEG-600-based precipitation. C) Amplification strategy, where protocol A corresponds to PCR-SISPA and protocol A.2 to MDA. D) Library preparation kit, where protocol A corresponds to the DNA Prep (M) Tagmentation kit and protocol A.3 to the TruSeq Nano DNA Library Prep kit. E) DNA/RNA extraction, where protocol A corresponds to the QIAamp Viral RNA Mini kit, protocol A.4 to the PureLink Viral RNA/DNA Mini kit and protocol A.4.1 to PureLink Viral RNA/DNA Mini kit followed by the cDNA second-strand synthesis. F) Boxplots showing the genome composition for the protocols. The central line represents the median, box limits indicate the Q1 and Q3 quartiles, and whiskers extend to Q1-1.5 IQR and Q3+1.5 IQR, marking the lowest and highest non-outlier values.

**Figure S3:** Composition of viruses by genome type for the protocols tested, excluding the *Microviridae* reads. A) Boxplots showing the genome composition excluding *Microviridae* reads. The central line represents the median, box limits indicate the Q1 and Q3 quartiles, and whiskers extend to Q1-1.5 IQR and Q3+1.5 IQR, marking the lowest and highest non-outlier values. B) Mean and 95% confidence interval (CI) of the log2 fold-change in each genome type after excluding *Microviridae* reads. Each modified protocol was compared to protocol A within its respective set of processed samples.

**Figure S4:** Structural and genomic characteristics of viruses identified by each protocol. Boxplots for each protocol showing the characteristics of viruses: circular or linear genome, enveloped or non enveloped structure, genome size (<15, 15-50 and >50 kb), viral particle size (<=50, 50-200 and >200 nm), and the proportion of phage-assigned particles. The central line represents the median, box limits indicate Q1 and Q3, and whiskers extend to the lowest and highest non-outlier values (Q1-1.5 IQR and Q3+1.5 IQR).

