## Supplementary material for "A Comparison of Methods for the Optimal Recovery of the Human Fecal Virome"

| | | Fastp | Human Read Removal | |
|---|---|---|---|---|
| **file** | **Raw Reads (R1)** | **Reads After** | **Reads Removed** | **Reads After** |
| 9092-U-virome_S11_L001_R1.fastq | 1664460 | 1417212 | 995 | 1416217 |
| 9092-1Q-virome_S3_L001_R1.fastq | 1742277 | 1419592 | 3481 | 1416111 |
| 9092-2Z-virome_S1_L001_R1.fastq | 1456005 | 1181383 | 175 | 1181208 |
| 2820-U-virome_S14_L001_R1.fastq | 1691399 | 1447463 | 2336 | 1445127 |
| 2820-2Q-virome_S8_L001_R1.fastq | 1460188 | 1261848 | 96 | 1261752 |
| 2820-2Z-virome_S2_L001_R1.fastq | 1697036 | 1510265 | 76 | 1510189 |
| 9324-U-virome_S12_L001_R1.fastq | 1600505 | 1213434 | 17666 | 1195768 |
| 9324-2Q-virome_S5_L001_R1.fastq | 1542093 | 1396271 | 91 | 1396180 |
| 9324-2Z-virome_S4_L001_R1.fastq | 1375155 | 1191969 | 1565 | 1190404 |
| 0168-U-virome_S13_L001_R1.fastq | 1538372 | 1329715 | 3111 | 1326604 |
| 0168-1Q-virome_S7_L001_R1.fastq | 1499517 | 1297634 | 1343 | 1296291 |
| 0168-1Z-virome_S6_L001_R1.fastq | 1862747 | 1591820 | 2930 | 1588890 |
| 8466-U-virome_S17_L001_R1.fastq | 1555191 | 1356565 | 322 | 1356243 |
| 8466-1Q-virome_S10_L001_R1.fastq | 1673012 | 1367067 | 255 | 1366812 |
| 8466-2Z-virome_S9_L001_R1.fastq | 1410253 | 1237346 | 767 | 1236579 |
| SAC1-viromeR_S55_L001_R1.fastq | 365571 | 331725 | 12 | 331713 |
| SAC2-viromeR_S56_L001_R1.fastq | 368602 | 337085 | 11 | 337074 |
| SBC1-virome_R1.fastq | 307751 | 283223 | 112 | 283111 |
| SBC2-virome_R1.fastq | 321036 | 293170 | 18 | 293152 |
| SAD1-viromeR_S61_L001_R1.fastq | 227262 | 213851 | 4 | 213847 |
| SAD2-virome_R1.fastq | 260881 | 244483 | 11 | 244472 |
| S1-5393-VIROME_S1_L001_R1.fastq | 5131783 | 4617437 | 3105 | 4614332 |
| S1-5393-viromeTruSeq.fastq | 1654933 | 1624596 | 1002 | 1623594 |
| S3-9504-VIROME_S3_L001_R1.fastq | 4781559 | 4434669 | 2810 | 4431859 |
| S3-9504-viromeTruSeq.fastq | 1217574 | 1021570 | 685 | 1020885 |
| S4-1876-VIROME_S4_L001_R1.fastq | 4913300 | 4540936 | 316 | 4540620 |
| S4-1876-viromeTruSeq.fastq | 1694898 | 1639087 | 112 | 1638975 |
| 14-2820AC-VIROME_S5_L001_R1.fastq | 3608775 | 3218914 | 3015 | 3215899 |

| | | | | |
|---|---|---|---|---|
| 14-2820AC-viromeTruSeq.fastq | 1390669 | 1333453 | 335 | 1333118 |
| 15-8466AC-VIROME_S6_L001_R1.fastq | 3811556 | 3541076 | 423 | 3540653 |
| 15-8466AC-viromeTruSeq.fastq | 1350120 | 1321047 | 113 | 1320934 |
| P1-2635-I-virome_R1.fastq | 897795 | 739396 | 39988 | 699408 |
| P1-2635-Q-virome_R1.fastq | 1020060 | 856378 | 3949 | 852429 |
| P1-2635-Ids-virome_R1.fastq | 651458 | 489337 | 48254 | 441083 |
| P2-6080-I-virome_R1.fastq | 919576 | 834110 | 2049 | 832061 |
| P2-6080-Q-virome_R1.fastq | 1018526 | 904308 | 414 | 903894 |
| P2-6080-Ids-virome_R1.fastq | 830553 | 744404 | 4126 | 740278 |
| P3-5566-I-virome_R1.fastq | 869113 | 791244 | 924 | 790320 |
| P3-5566-Q-virome_R1.fastq | 895461 | 804130 | 770 | 803360 |
| P3-5566-Ids-virome_R1.fastq | 1018301 | 916744 | 2171 | 914573 |
| P4-1728-I-virome_R1.fastq | 999725 | 908923 | 786 | 908137 |
| P4-1728-Q-virome_R1.fastq | 969928 | 880027 | 950 | 879077 |
| P4-1728-Ids-virome_R1.fastq | 983342 | 906062 | 1385 | 904677 |
| P5-8986-I-virome_R1.fastq | 758798 | 693210 | 3668 | 689542 |
| P5-8986-Q-virome_R1.fastq | 891602 | 821949 | 2757 | 819192 |
| P5-8986-Ids-virome_R1.fastq | 903287 | 815061 | 11596 | 803465 |
| P6-8992-I-virome_R1.fastq | 873992 | 789156 | 787 | 788369 |
| P6-8992-Q-virome_R1.fastq | 746119 | 681266 | 599 | 680667 |
| P6-8992-Ids-virome_R1.fastq | 1103322 | 987145 | 1921 | 985224 |

| file | format | type | Reads After Fastp | sum_len | min_len | avg_len | max_len |
|---|---|---|---|---|---|---|---|
| 9092-U-virome_S11_L001_R1.fastq | FASTQ | DNA | 1417212 | 150912234 | 50 | 106.5 | 151 |
| 9092-U-virome_S11_L001_R2.fastq | FASTQ | DNA | 1417212 | 150612398 | 50 | 106.3 | 151 |
| 9092-1Q-virome_S3_L001_R1.fastq | FASTQ | DNA | 1419592 | 135065033 | 50 | 95.1 | 151 |
| 9092-1Q-virome_S3_L001_R2.fastq | FASTQ | DNA | 1419592 | 134676676 | 50 | 94.9 | 151 |
| 9092-2Z-virome_S1_L001_R1.fastq | FASTQ | DNA | 1181383 | 117247142 | 50 | 99.2 | 151 |
| 9092-2Z-virome_S1_L001_R2.fastq | FASTQ | DNA | 1181383 | 116990968 | 50 | 99 | 151 |
| 2820-U-virome_S14_L001_R1.fastq | FASTQ | DNA | 1447463 | 158570239 | 50 | 109.6 | 151 |
| 2820-U-virome_S14_L001_R2.fastq | FASTQ | DNA | 1447463 | 158298344 | 50 | 109.4 | 151 |
| 2820-2Q-virome_S8_L001_R1.fastq | FASTQ | DNA | 1261848 | 124280606 | 50 | 98.5 | 151 |
| 2820-2Q-virome_S8_L001_R2.fastq | FASTQ | DNA | 1261848 | 123946963 | 50 | 98.2 | 151 |
| 2820-2Z-virome_S2_L001_R1.fastq | FASTQ | DNA | 1510265 | 165383692 | 50 | 109.5 | 151 |
| 2820-2Z-virome_S2_L001_R2.fastq | FASTQ | DNA | 1510265 | 165161416 | 50 | 109.4 | 151 |
| 9324-U-virome_S12_L001_R1.fastq | FASTQ | DNA | 1213434 | 121680025 | 50 | 100.3 | 151 |
| 9324-U-virome_S12_L001_R2.fastq | FASTQ | DNA | 1213434 | 121315226 | 50 | 100 | 151 |
| 9324-2Q-virome_S5_L001_R1.fastq | FASTQ | DNA | 1396271 | 155501595 | 50 | 111.4 | 151 |
| 9324-2Q-virome_S5_L001_R2.fastq | FASTQ | DNA | 1396271 | 155197498 | 50 | 111.2 | 151 |
| 9324-2Z-virome_S4_L001_R1.fastq | FASTQ | DNA | 1191969 | 127427880 | 50 | 106.9 | 151 |
| 9324-2Z-virome_S4_L001_R2.fastq | FASTQ | DNA | 1191969 | 127112831 | 50 | 106.6 | 151 |
| 0168-U-virome_S13_L001_R1.fastq | FASTQ | DNA | 1329715 | 145641658 | 50 | 109.5 | 151 |
| 0168-U-virome_S13_L001_R2.fastq | FASTQ | DNA | 1329715 | 145356512 | 50 | 109.3 | 151 |
| 0168-1Q-virome_S7_L001_R1.fastq | FASTQ | DNA | 1297634 | 140486614 | 50 | 108.3 | 151 |
| 0168-1Q-virome_S7_L001_R2.fastq | FASTQ | DNA | 1297634 | 140179597 | 50 | 108 | 151 |
| 0168-1Z-virome_S6_L001_R1.fastq | FASTQ | DNA | 1591820 | 163870803 | 50 | 102.9 | 151 |
| 0168-1Z-virome_S6_L001_R2.fastq | FASTQ | DNA | 1591820 | 163539449 | 50 | 102.7 | 151 |
| 8466-U-virome_S17_L001_R1.fastq | FASTQ | DNA | 1356565 | 152210113 | 50 | 112.2 | 151 |
| 8466-U-virome_S17_L001_R2.fastq | FASTQ | DNA | 1356565 | 151986613 | 50 | 112 | 151 |
| 8466-1Q-virome_S10_L001_R1.fastq | FASTQ | DNA | 1367067 | 132809156 | 50 | 97.1 | 151 |
| 8466-1Q-virome_S10_L001_R2.fastq | FASTQ | DNA | 1367067 | 132435012 | 50 | 96.9 | 151 |
| 8466-2Z-virome_S9_L001_R1.fastq | FASTQ | DNA | 1237346 | 131272792 | 50 | 106.1 | 151 |

| | | | | | | | |
|---|---|---|---|---|---|---|---|
| 8466-2Z-virome_S9_L001_R2.fastq | FASTQ | DNA | 1237346 | 130968555 | 50 | 105.8 | 151 |
| SAC1-viromeR_S55_L001_R1.fastq | FASTQ | DNA | 331725 | 39031581 | 50 | 117.7 | 151 |
| SAC1-viromeR_S55_L001_R2.fastq | FASTQ | DNA | 331725 | 38974631 | 50 | 117.5 | 151 |
| SAC2-viromeR_S56_L001_R1.fastq | FASTQ | DNA | 337085 | 40042025 | 50 | 118.8 | 151 |
| SAC2-viromeR_S56_L001_R2.fastq | FASTQ | DNA | 337085 | 39984546 | 50 | 118.6 | 151 |
| SBC1-virome_R1.fastq | FASTQ | DNA | 283223 | 33736104 | 50 | 119.1 | 151 |
| SBC1-virome_R2.fastq | FASTQ | DNA | 283223 | 33692234 | 50 | 119 | 151 |
| SBC2-virome_R1.fastq | FASTQ | DNA | 293170 | 34506971 | 50 | 117.7 | 151 |
| SBC2-virome_R2.fastq | FASTQ | DNA | 293170 | 34461852 | 50 | 117.5 | 151 |
| SAD1-viromeR_S61_L001_R1.fastq | FASTQ | DNA | 213851 | 27269445 | 50 | 127.5 | 151 |
| SAD1-viromeR_S61_L001_R2.fastq | FASTQ | DNA | 213851 | 27236344 | 50 | 127.4 | 151 |
| SAD2-virome_R1.fastq | FASTQ | DNA | 244483 | 31470425 | 50 | 128.7 | 151 |
| SAD2-virome_R2.fastq | FASTQ | DNA | 244483 | 31440229 | 50 | 128.6 | 151 |
| S1-5393-VIROME_S1_L001_R1.fastq | FASTQ | DNA | 4617437 | 539934391 | 50 | 116.9 | 151 |
| S1-5393-VIROME_S1_L001_R2.fastq | FASTQ | DNA | 4617437 | 539152616 | 50 | 116.8 | 151 |
| S1-5393-viromeTruSeq.fastq | FASTQ | DNA | 1624596 | 237353678 | 50 | 146.1 | 151 |
| S3-9504-VIROME_S3_L001_R1.fastq | FASTQ | DNA | 4434669 | 539628515 | 50 | 121.7 | 151 |
| S3-9504-VIROME_S3_L001_R2.fastq | FASTQ | DNA | 4434669 | 539073611 | 50 | 121.6 | 151 |
| S3-9504-viromeTruSeq.fastq | FASTQ | DNA | 1021570 | 132551922 | 50 | 129.8 | 151 |
| S4-1876-VIROME_S4_L001_R1.fastq | FASTQ | DNA | 4540936 | 557296874 | 50 | 122.7 | 151 |
| S4-1876-VIROME_S4_L001_R2.fastq | FASTQ | DNA | 4540936 | 556785316 | 50 | 122.6 | 151 |
| S4-1876-viromeTruSeq.fastq | FASTQ | DNA | 1639087 | 234162235 | 50 | 142.9 | 151 |
| 14-2820AC-VIROME_S5_L001_R1.fastq | FASTQ | DNA | 3218914 | 379551536 | 50 | 117.9 | 151 |
| 14-2820AC-VIROME_S5_L001_R2.fastq | FASTQ | DNA | 3218914 | 379232324 | 50 | 117.8 | 151 |
| 14-2820AC-viromeTruSeq.fastq | FASTQ | DNA | 1333453 | 188490601 | 50 | 141.4 | 151 |
| 15-8466AC-VIROME_S6_L001_R1.fastq | FASTQ | DNA | 3541076 | 443647313 | 50 | 125.3 | 151 |
| 15-8466AC-VIROME_S6_L001_R2.fastq | FASTQ | DNA | 3541076 | 443317296 | 50 | 125.2 | 151 |
| 15-8466AC-viromeTruSeq.fastq | FASTQ | DNA | 1321047 | 192023188 | 50 | 145.4 | 151 |
| P1-2635-I-virome_R1.fastq | FASTQ | DNA | 739396 | 83607983 | 50 | 113.1 | 151 |
| P1-2635-I-virome_R2.fastq | FASTQ | DNA | 739396 | 83400085 | 50 | 112.8 | 151 |

| | | | | | | | |
|---|---|---|---|---|---|---|---|
| P1-2635-Q-virome_R1.fastq | FASTQ | DNA | 856378 | 90701946 | 50 | 105.9 | 151 |
| P1-2635-Q-virome_R2.fastq | FASTQ | DNA | 856378 | 90521025 | 50 | 105.7 | 151 |
| P1-2635-Ids-virome_R1.fastq | FASTQ | DNA | 489337 | 55140137 | 50 | 112.7 | 151 |
| P1-2635-Ids-virome_R2.fastq | FASTQ | DNA | 489337 | 54935787 | 50 | 112.3 | 151 |
| P2-6080-I-virome_R1.fastq | FASTQ | DNA | 834110 | 100477574 | 50 | 120.5 | 151 |
| P2-6080-I-virome_R2.fastq | FASTQ | DNA | 834110 | 100402084 | 50 | 120.4 | 151 |
| P2-6080-Q-virome_R1.fastq | FASTQ | DNA | 904308 | 103042428 | 50 | 113.9 | 151 |
| P2-6080-Q-virome_R2.fastq | FASTQ | DNA | 904308 | 102909497 | 50 | 113.8 | 151 |
| P2-6080-Ids-virome_R1.fastq | FASTQ | DNA | 744404 | 90430974 | 50 | 121.5 | 151 |
| P2-6080-Ids-virome_R2.fastq | FASTQ | DNA | 744404 | 90359527 | 50 | 121.4 | 151 |
| P3-5566-I-virome_R1.fastq | FASTQ | DNA | 791244 | 95411484 | 50 | 120.6 | 151 |
| P3-5566-I-virome_R2.fastq | FASTQ | DNA | 791244 | 95306897 | 50 | 120.5 | 151 |
| P3-5566-Q-virome_R1.fastq | FASTQ | DNA | 804130 | 94598451 | 50 | 117.6 | 151 |
| P3-5566-Q-virome_R2.fastq | FASTQ | DNA | 804130 | 94476132 | 50 | 117.5 | 151 |
| P3-5566-Ids-virome_R1.fastq | FASTQ | DNA | 916744 | 111275067 | 50 | 121.4 | 151 |
| P3-5566-Ids-virome_R2.fastq | FASTQ | DNA | 916744 | 111184608 | 50 | 121.3 | 151 |
| P4-1728-I-virome_R1.fastq | FASTQ | DNA | 908923 | 108693950 | 50 | 119.6 | 151 |
| P4-1728-I-virome_R2.fastq | FASTQ | DNA | 908923 | 108614694 | 50 | 119.5 | 151 |
| P4-1728-Q-virome_R1.fastq | FASTQ | DNA | 880027 | 99717849 | 50 | 113.3 | 151 |
| P4-1728-Q-virome_R2.fastq | FASTQ | DNA | 880027 | 99584378 | 50 | 113.2 | 151 |
| P4-1728-Ids-virome_R1.fastq | FASTQ | DNA | 906062 | 109312118 | 50 | 120.6 | 151 |
| P4-1728-Ids-virome_R2.fastq | FASTQ | DNA | 906062 | 109226587 | 50 | 120.6 | 151 |
| P5-8986-I-virome_R1.fastq | FASTQ | DNA | 693210 | 83924257 | 50 | 121.1 | 151 |
| P5-8986-I-virome_R2.fastq | FASTQ | DNA | 693210 | 83841291 | 50 | 120.9 | 151 |
| P5-8986-Q-virome_R1.fastq | FASTQ | DNA | 821949 | 101157525 | 50 | 123.1 | 151 |
| P5-8986-Q-virome_R2.fastq | FASTQ | DNA | 821949 | 101073179 | 50 | 123 | 151 |
| P5-8986-Ids-virome_R1.fastq | FASTQ | DNA | 815061 | 97085018 | 50 | 119.1 | 151 |
| P5-8986-Ids-virome_R2.fastq | FASTQ | DNA | 815061 | 96956914 | 50 | 119 | 151 |
| P6-8992-I-virome_R1.fastq | FASTQ | DNA | 789156 | 94368196 | 50 | 119.6 | 151 |
| P6-8992-I-virome_R2.fastq | FASTQ | DNA | 789156 | 94291411 | 50 | 119.5 | 151 |

| | | | | | | | |
|---|---|---|---|---|---|---|---|
| P6-8992-Q-virome_R1.fastq | FASTQ | DNA | 681266 | 82094810 | 50 | 120.5 | 151 |
| P6-8992-Q-virome_R2.fastq | FASTQ | DNA | 681266 | 82024261 | 50 | 120.4 | 151 |
| P6-8992-Ids-virome_R1.fastq | FASTQ | DNA | 987145 | 117667768 | 50 | 119.2 | 151 |
| P6-8992-Ids-virome_R2.fastq | FASTQ | DNA | 987145 | 117572718 | 50 | 119.1 | 151 |

| file | format | type | num_seqs | sum_len | min_len | avg_len | max_len |
|---|---|---|---|---|---|---|---|
| 9092-U-virome_S11_L001_R1.fastq | FASTQ | DNA | 1416217 | 150818638 | 50 | 106.5 | 151 |
| 9092-U-virome_S11_L001_R2.fastq | FASTQ | DNA | 1416217 | 150518841 | 50 | 106.3 | 151 |
| 9092-1Q-virome_S3_L001_R1.fastq | FASTQ | DNA | 1416111 | 134805045 | 50 | 95.2 | 151 |
| 9092-1Q-virome_S3_L001_R2.fastq | FASTQ | DNA | 1416111 | 134440192 | 50 | 94.9 | 151 |
| 9092-2Z-virome_S1_L001_R1.fastq | FASTQ | DNA | 1181208 | 117230893 | 50 | 99.2 | 151 |
| 9092-2Z-virome_S1_L001_R2.fastq | FASTQ | DNA | 1181208 | 116974739 | 50 | 99 | 151 |
| 2820-U-virome_S14_L001_R1.fastq | FASTQ | DNA | 1445127 | 158420790 | 50 | 109.6 | 151 |
| 2820-U-virome_S14_L001_R2.fastq | FASTQ | DNA | 1445127 | 158149002 | 50 | 109.4 | 151 |
| 2820-2Q-virome_S8_L001_R1.fastq | FASTQ | DNA | 1261752 | 124271931 | 50 | 98.5 | 151 |
| 2820-2Q-virome_S8_L001_R2.fastq | FASTQ | DNA | 1261752 | 123938317 | 50 | 98.2 | 151 |
| 2820-2Z-virome_S2_L001_R1.fastq | FASTQ | DNA | 1510189 | 165376165 | 50 | 109.5 | 151 |
| 2820-2Z-virome_S2_L001_R2.fastq | FASTQ | DNA | 1510189 | 165153886 | 50 | 109.4 | 151 |
| 9324-U-virome_S12_L001_R1.fastq | FASTQ | DNA | 1195768 | 120498045 | 50 | 100.8 | 151 |
| 9324-U-virome_S12_L001_R2.fastq | FASTQ | DNA | 1195768 | 120136378 | 50 | 100.5 | 151 |
| 9324-2Q-virome_S5_L001_R1.fastq | FASTQ | DNA | 1396180 | 155492855 | 50 | 111.4 | 151 |
| 9324-2Q-virome_S5_L001_R2.fastq | FASTQ | DNA | 1396180 | 155188750 | 50 | 111.2 | 151 |
| 9324-2Z-virome_S4_L001_R1.fastq | FASTQ | DNA | 1190404 | 127267155 | 50 | 106.9 | 151 |
| 9324-2Z-virome_S4_L001_R2.fastq | FASTQ | DNA | 1190404 | 126952209 | 50 | 106.6 | 151 |
| 0168-U-virome_S13_L001_R1.fastq | FASTQ | DNA | 1326604 | 145438214 | 50 | 109.6 | 151 |
| 0168-U-virome_S13_L001_R2.fastq | FASTQ | DNA | 1326604 | 145153535 | 50 | 109.4 | 151 |
| 0168-1Q-virome_S7_L001_R1.fastq | FASTQ | DNA | 1296291 | 140361276 | 50 | 108.3 | 151 |
| 0168-1Q-virome_S7_L001_R2.fastq | FASTQ | DNA | 1296291 | 140054325 | 50 | 108 | 151 |
| 0168-1Z-virome_S6_L001_R1.fastq | FASTQ | DNA | 1588890 | 163567631 | 50 | 102.9 | 151 |
| 0168-1Z-virome_S6_L001_R2.fastq | FASTQ | DNA | 1588890 | 163236313 | 50 | 102.7 | 151 |
| 8466-U-virome_S17_L001_R1.fastq | FASTQ | DNA | 1356243 | 152190019 | 50 | 112.2 | 151 |
| 8466-U-virome_S17_L001_R2.fastq | FASTQ | DNA | 1356243 | 151966594 | 50 | 112 | 151 |
| 8466-1Q-virome_S10_L001_R1.fastq | FASTQ | DNA | 1366812 | 132788288 | 50 | 97.2 | 151 |
| 8466-1Q-virome_S10_L001_R2.fastq | FASTQ | DNA | 1366812 | 132414153 | 50 | 96.9 | 151 |
| 8466-2Z-virome_S9_L001_R1.fastq | FASTQ | DNA | 1236579 | 131197456 | 50 | 106.1 | 151 |

| | | | | | | | |
|---|---|---|---|---|---|---|---|
| 8466-2Z-virome_S9_L001_R2.fastq | FASTQ | DNA | 1236579 | 130893437 | 50 | 105.9 | 151 |
| SAC1-viromeR_S55_L001_R1.fastq | FASTQ | DNA | 331713 | 39030840 | 50 | 117.7 | 151 |
| SAC1-viromeR_S55_L001_R2.fastq | FASTQ | DNA | 331713 | 38973890 | 50 | 117.5 | 151 |
| SAC2-viromeR_S56_L001_R1.fastq | FASTQ | DNA | 337074 | 40041317 | 50 | 118.8 | 151 |
| SAC2-viromeR_S56_L001_R2.fastq | FASTQ | DNA | 337074 | 39983839 | 50 | 118.6 | 151 |
| SBC1-virome_R1.fastq | FASTQ | DNA | 283111 | 33723241 | 50 | 119.1 | 151 |
| SBC1-virome_R2.fastq | FASTQ | DNA | 283111 | 33679378 | 50 | 119 | 151 |
| SBC2-virome_R1.fastq | FASTQ | DNA | 293152 | 34505773 | 50 | 117.7 | 151 |
| SBC2-virome_R2.fastq | FASTQ | DNA | 293152 | 34460656 | 50 | 117.6 | 151 |
| SAD1-viromeR_S61_L001_R1.fastq | FASTQ | DNA | 213847 | 27268918 | 50 | 127.5 | 151 |
| SAD1-viromeR_S61_L001_R2.fastq | FASTQ | DNA | 213847 | 27235818 | 50 | 127.4 | 151 |
| SAD2-virome_R1.fastq | FASTQ | DNA | 244472 | 31469024 | 50 | 128.7 | 151 |
| SAD2-virome_R2.fastq | FASTQ | DNA | 244472 | 31438830 | 50 | 128.6 | 151 |
| S1-5393-VIROME_S1_L001_R1.fastq | FASTQ | DNA | 4614332 | 539714771 | 50 | 117 | 151 |
| S1-5393-VIROME_S1_L001_R2.fastq | FASTQ | DNA | 4614332 | 538933963 | 50 | 116.8 | 151 |
| S1-5393-viromeTruSeq.fastq | FASTQ | DNA | 1623594 | 237215689 | 50 | 146.1 | 151 |
| S3-9504-VIROME_S3_L001_R1.fastq | FASTQ | DNA | 4431859 | 539433655 | 50 | 121.7 | 151 |
| S3-9504-VIROME_S3_L001_R2.fastq | FASTQ | DNA | 4431859 | 538880314 | 50 | 121.6 | 151 |
| S3-9504-viromeTruSeq.fastq | FASTQ | DNA | 1020885 | 132461922 | 50 | 129.8 | 151 |
| S4-1876-VIROME_S4_L001_R1.fastq | FASTQ | DNA | 4540620 | 557276227 | 50 | 122.7 | 151 |
| S4-1876-VIROME_S4_L001_R2.fastq | FASTQ | DNA | 4540620 | 556764782 | 50 | 122.6 | 151 |
| S4-1876-viromeTruSeq.fastq | FASTQ | DNA | 1638975 | 234150063 | 50 | 142.9 | 151 |
| 14-2820AC-VIROME_S5_L001_R1.fas | FASTQ | DNA | 3215899 | 379364407 | 50 | 118 | 151 |
| 14-2820AC-VIROME_S5_L001_R2.fas | FASTQ | DNA | 3215899 | 379045258 | 50 | 117.9 | 151 |
| 14-2820AC-viromeTruSeq.fastq | FASTQ | DNA | 1333118 | 188451951 | 50 | 141.4 | 151 |
| 15-8466AC-VIROME_S6_L001_R1.fas | FASTQ | DNA | 3540653 | 443602853 | 50 | 125.3 | 151 |
| 15-8466AC-VIROME_S6_L001_R2.fas | FASTQ | DNA | 3540653 | 443272736 | 50 | 125.2 | 151 |
| 15-8466AC-viromeTruSeq.fastq | FASTQ | DNA | 1320934 | 192010897 | 50 | 145.4 | 151 |
| P1-2635-I-virome_R1.fastq | FASTQ | DNA | 699408 | 80714433 | 50 | 115.4 | 151 |
| P1-2635-I-virome_R2.fastq | FASTQ | DNA | 699408 | 80524211 | 50 | 115.1 | 151 |

| | | | | | | | |
|---|---|---|---|---|---|---|---|
| P1-2635-Q-virome_R1.fastq | FASTQ | DNA | 852429 | 90432867 | 50 | 106.1 | 151 |
| P1-2635-Q-virome_R2.fastq | FASTQ | DNA | 852429 | 90253161 | 50 | 105.9 | 151 |
| P1-2635-Ids-virome_R1.fastq | FASTQ | DNA | 441083 | 51649165 | 50 | 117.1 | 151 |
| P1-2635-Ids-virome_R2.fastq | FASTQ | DNA | 441083 | 51471458 | 50 | 116.7 | 151 |
| P2-6080-I-virome_R1.fastq | FASTQ | DNA | 832061 | 100333759 | 50 | 120.6 | 151 |
| P2-6080-I-virome_R2.fastq | FASTQ | DNA | 832061 | 100258642 | 50 | 120.5 | 151 |
| P2-6080-Q-virome_R1.fastq | FASTQ | DNA | 903894 | 103014545 | 50 | 114 | 151 |
| P2-6080-Q-virome_R2.fastq | FASTQ | DNA | 903894 | 102881755 | 50 | 113.8 | 151 |
| P2-6080-Ids-virome_R1.fastq | FASTQ | DNA | 740278 | 90141001 | 50 | 121.8 | 151 |
| P2-6080-Ids-virome_R2.fastq | FASTQ | DNA | 740278 | 90071428 | 50 | 121.7 | 151 |
| P3-5566-I-virome_R1.fastq | FASTQ | DNA | 790320 | 95346188 | 50 | 120.6 | 151 |
| P3-5566-I-virome_R2.fastq | FASTQ | DNA | 790320 | 95242136 | 50 | 120.5 | 151 |
| P3-5566-Q-virome_R1.fastq | FASTQ | DNA | 803360 | 94546066 | 50 | 117.7 | 151 |
| P3-5566-Q-virome_R2.fastq | FASTQ | DNA | 803360 | 94424009 | 50 | 117.5 | 151 |
| P3-5566-Ids-virome_R1.fastq | FASTQ | DNA | 914573 | 111123518 | 50 | 121.5 | 151 |
| P3-5566-Ids-virome_R2.fastq | FASTQ | DNA | 914573 | 111033843 | 50 | 121.4 | 151 |
| P4-1728-I-virome_R1.fastq | FASTQ | DNA | 908137 | 108638426 | 50 | 119.6 | 151 |
| P4-1728-I-virome_R2.fastq | FASTQ | DNA | 908137 | 108559729 | 50 | 119.5 | 151 |
| P4-1728-Q-virome_R1.fastq | FASTQ | DNA | 879077 | 99651340 | 50 | 113.4 | 151 |
| P4-1728-Q-virome_R2.fastq | FASTQ | DNA | 879077 | 99518234 | 50 | 113.2 | 151 |
| P4-1728-Ids-virome_R1.fastq | FASTQ | DNA | 904677 | 109211754 | 50 | 120.7 | 151 |
| P4-1728-Ids-virome_R2.fastq | FASTQ | DNA | 904677 | 109126842 | 50 | 120.6 | 151 |
| P5-8986-I-virome_R1.fastq | FASTQ | DNA | 689542 | 83661825 | 50 | 121.3 | 151 |
| P5-8986-I-virome_R2.fastq | FASTQ | DNA | 689542 | 83580389 | 50 | 121.2 | 151 |
| P5-8986-Q-virome_R1.fastq | FASTQ | DNA | 819192 | 100960457 | 50 | 123.2 | 151 |
| P5-8986-Q-virome_R2.fastq | FASTQ | DNA | 819192 | 100878043 | 50 | 123.1 | 151 |
| P5-8986-Ids-virome_R1.fastq | FASTQ | DNA | 803465 | 96233288 | 50 | 119.8 | 151 |
| P5-8986-Ids-virome_R2.fastq | FASTQ | DNA | 803465 | 96115897 | 50 | 119.6 | 151 |
| P6-8992-I-virome_R1.fastq | FASTQ | DNA | 788369 | 94310985 | 50 | 119.6 | 151 |
| P6-8992-I-virome_R2.fastq | FASTQ | DNA | 788369 | 94234601 | 50 | 119.5 | 151 |

| | | | | | | | |
|---|---|---|---|---|---|---|---|
| P6-8992-Q-virome_R1.fastq | FASTQ | DNA | 680667 | 82053389 | 50 | 120.5 | 151 |
| P6-8992-Q-virome_R2.fastq | FASTQ | DNA | 680667 | 81983306 | 50 | 120.4 | 151 |
| P6-8992-Ids-virome_R1.fastq | FASTQ | DNA | 985224 | 117530364 | 50 | 119.3 | 151 |
| P6-8992-Ids-virome_R2.fastq | FASTQ | DNA | 985224 | 117436544 | 50 | 119.2 | 151 |

| file | format | type | num_seqs | sum_len | min_len | avg_len | max_len |
|---|---|---|---|---|---|---|---|
| 9092-U-virome_S11_L001_R1.fastq | FASTQ | DNA | 1664460 | 163186954 | 35 | 98 | 151 |
| 9092-U-virome_S11_L001_R2.fastq | FASTQ | DNA | 1664460 | 163495575 | 35 | 98.2 | 151 |
| 9092-1Q-virome_S3_L001_R1.fastq | FASTQ | DNA | 1742277 | 151362771 | 35 | 86.9 | 151 |
| 9092-1Q-virome_S3_L001_R2.fastq | FASTQ | DNA | 1742277 | 151947123 | 35 | 87.2 | 151 |
| 9092-2Z-virome_S1_L001_R1.fastq | FASTQ | DNA | 1456005 | 131401478 | 35 | 90.2 | 151 |
| 9092-2Z-virome_S1_L001_R2.fastq | FASTQ | DNA | 1456005 | 131680471 | 35 | 90.4 | 151 |
| 2820-U-virome_S14_L001_R1.fastq | FASTQ | DNA | 1691399 | 171695369 | 35 | 101.5 | 151 |
| 2820-U-virome_S14_L001_R2.fastq | FASTQ | DNA | 1691399 | 171895716 | 35 | 101.6 | 151 |
| 2820-2Q-virome_S8_L001_R1.fastq | FASTQ | DNA | 1460188 | 134566658 | 35 | 92.2 | 151 |
| 2820-2Q-virome_S8_L001_R2.fastq | FASTQ | DNA | 1460188 | 135030567 | 35 | 92.5 | 151 |
| 2820-2Z-virome_S2_L001_R1.fastq | FASTQ | DNA | 1697036 | 174879989 | 35 | 103.1 | 151 |
| 2820-2Z-virome_S2_L001_R2.fastq | FASTQ | DNA | 1697036 | 175193919 | 35 | 103.2 | 151 |
| 9324-U-virome_S12_L001_R1.fastq | FASTQ | DNA | 1600505 | 148133815 | 35 | 92.6 | 151 |
| 9324-U-virome_S12_L001_R2.fastq | FASTQ | DNA | 1600505 | 148572861 | 35 | 92.8 | 151 |
| 9324-2Q-virome_S5_L001_R1.fastq | FASTQ | DNA | 1542093 | 163786013 | 35 | 106.2 | 151 |
| 9324-2Q-virome_S5_L001_R2.fastq | FASTQ | DNA | 1542093 | 164411454 | 35 | 106.6 | 151 |
| 9324-2Z-virome_S4_L001_R1.fastq | FASTQ | DNA | 1375155 | 137902445 | 35 | 100.3 | 151 |
| 9324-2Z-virome_S4_L001_R2.fastq | FASTQ | DNA | 1375155 | 138305700 | 35 | 100.6 | 151 |
| 0168-U-virome_S13_L001_R1.fastq | FASTQ | DNA | 1538372 | 157572702 | 35 | 102.4 | 151 |
| 0168-U-virome_S13_L001_R2.fastq | FASTQ | DNA | 1538372 | 157771802 | 35 | 102.6 | 151 |
| 0168-1Q-virome_S7_L001_R1.fastq | FASTQ | DNA | 1499517 | 151049673 | 35 | 100.7 | 151 |
| 0168-1Q-virome_S7_L001_R2.fastq | FASTQ | DNA | 1499517 | 151305475 | 35 | 100.9 | 151 |
| 0168-1Z-virome_S6_L001_R1.fastq | FASTQ | DNA | 1862747 | 177209501 | 35 | 95.1 | 151 |
| 0168-1Z-virome_S6_L001_R2.fastq | FASTQ | DNA | 1862747 | 177469015 | 35 | 95.3 | 151 |
| 8466-U-virome_S17_L001_R1.fastq | FASTQ | DNA | 1555191 | 163086310 | 35 | 104.9 | 151 |
| 8466-U-virome_S17_L001_R2.fastq | FASTQ | DNA | 1555191 | 163322109 | 35 | 105 | 151 |
| 8466-1Q-virome_S10_L001_R1.fastq | FASTQ | DNA | 1673012 | 147733332 | 35 | 88.3 | 151 |
| 8466-1Q-virome_S10_L001_R2.fastq | FASTQ | DNA | 1673012 | 148159065 | 35 | 88.6 | 151 |
| 8466-2Z-virome_S9_L001_R1.fastq | FASTQ | DNA | 1410253 | 140124332 | 35 | 99.4 | 151 |

| | | | | | | | |
|---|---|---|---|---|---|---|---|
| 8466-2Z-virome_S9_L001_R2.fastq | FASTQ | DNA | 1410253 | 140360043 | 35 | 99.5 | 151 |
| SAC1-viromeR_S55_L001_R1.fastq | FASTQ | DNA | 365571 | 40885137 | 35 | 111.8 | 151 |
| SAC1-viromeR_S55_L001_R2.fastq | FASTQ | DNA | 365571 | 40986018 | 35 | 112.1 | 151 |
| SAC2-viromeR_S56_L001_R1.fastq | FASTQ | DNA | 368602 | 41778370 | 35 | 113.3 | 151 |
| SAC2-viromeR_S56_L001_R2.fastq | FASTQ | DNA | 368602 | 41867666 | 35 | 113.6 | 151 |
| SBC1-virome_R1.fastq | FASTQ | DNA | 307751 | 35187033 | 35 | 114.3 | 151 |
| SBC1-virome_R2.fastq | FASTQ | DNA | 307751 | 35264133 | 35 | 114.6 | 151 |
| SBC2-virome_R1.fastq | FASTQ | DNA | 321036 | 36081076 | 35 | 112.4 | 151 |
| SBC2-virome_R2.fastq | FASTQ | DNA | 321036 | 36164267 | 35 | 112.6 | 151 |
| SAD1-viromeR_S61_L001_R1.fastq | FASTQ | DNA | 227262 | 28121635 | 35 | 123.7 | 151 |
| SAD1-viromeR_S61_L001_R2.fastq | FASTQ | DNA | 227262 | 28171300 | 35 | 124 | 151 |
| SAD2-virome_R1.fastq | FASTQ | DNA | 260881 | 32543242 | 35 | 124.7 | 151 |
| SAD2-virome_R2.fastq | FASTQ | DNA | 260881 | 32585940 | 35 | 124.9 | 151 |
| S1-5393-VIROME_S1_L001_R1.fastq | FASTQ | DNA | 5131783 | 570255202 | 35 | 111.1 | 151 |
| S1-5393-VIROME_S1_L001_R2.fastq | FASTQ | DNA | 5131783 | 571449917 | 35 | 111.4 | 151 |
| S1-5393-viromeTruSeq.fastq | FASTQ | DNA | 1654933 | 242120533 | 35 | 146.3 | 151 |
| S3-9504-VIROME_S3_L001_R1.fastq | FASTQ | DNA | 4781559 | 561014926 | 35 | 117.3 | 151 |
| S3-9504-VIROME_S3_L001_R2.fastq | FASTQ | DNA | 4781559 | 561767293 | 35 | 117.5 | 151 |
| S3-9504-viromeTruSeq.fastq | FASTQ | DNA | 1217574 | 161163452 | 35 | 132.4 | 151 |
| S4-1876-VIROME_S4_L001_R1.fastq | FASTQ | DNA | 4913300 | 578219441 | 35 | 117.7 | 151 |
| S4-1876-VIROME_S4_L001_R2.fastq | FASTQ | DNA | 4913300 | 578866901 | 35 | 117.8 | 151 |
| S4-1876-viromeTruSeq.fastq | FASTQ | DNA | 1694898 | 242602985 | 35 | 143.1 | 151 |
| 14-2820AC-VIROME_S5_L001_R1.fastq | FASTQ | DNA | 3608775 | 402953356 | 35 | 111.7 | 151 |
| 14-2820AC-VIROME_S5_L001_R2.fastq | FASTQ | DNA | 3608775 | 403287252 | 35 | 111.8 | 151 |
| 14-2820AC-viromeTruSeq.fastq | FASTQ | DNA | 1390669 | 197636203 | 35 | 142.1 | 151 |
| 15-8466AC-VIROME_S6_L001_R1.fastq | FASTQ | DNA | 3811556 | 460210413 | 35 | 120.7 | 151 |
| 15-8466AC-VIROME_S6_L001_R2.fastq | FASTQ | DNA | 3811556 | 460772911 | 35 | 120.9 | 151 |
| 15-8466AC-viromeTruSeq.fastq | FASTQ | DNA | 1350120 | 197017780 | 35 | 145.9 | 151 |
| P1-2635-I-virome_R1.fastq | FASTQ | DNA | 897795 | 99878387 | 35 | 111.2 | 151 |
| P1-2635-I-virome_R2.fastq | FASTQ | DNA | 897795 | 100125848 | 35 | 111.5 | 151 |

| | | | | | | | |
|---|---|---|---|---|---|---|---|
| P1-2635-Q-virome_R1.fastq | FASTQ | DNA | 1020060 | 100205640 | 35 | 98.2 | 151 |
| P1-2635-Q-virome_R2.fastq | FASTQ | DNA | 1020060 | 100449102 | 35 | 98.5 | 151 |
| P1-2635-Ids-virome_R1.fastq | FASTQ | DNA | 651458 | 73893996 | 35 | 113.4 | 151 |
| P1-2635-Ids-virome_R2.fastq | FASTQ | DNA | 651458 | 74219429 | 35 | 113.9 | 151 |
| P2-6080-I-virome_R1.fastq | FASTQ | DNA | 919576 | 105645784 | 35 | 114.9 | 151 |
| P2-6080-I-virome_R2.fastq | FASTQ | DNA | 919576 | 105752314 | 35 | 115 | 151 |
| P2-6080-Q-virome_R1.fastq | FASTQ | DNA | 1018526 | 109113824 | 35 | 107.1 | 151 |
| P2-6080-Q-virome_R2.fastq | FASTQ | DNA | 1018526 | 109229682 | 35 | 107.2 | 151 |
| P2-6080-Ids-virome_R1.fastq | FASTQ | DNA | 830553 | 96425469 | 35 | 116.1 | 151 |
| P2-6080-Ids-virome_R2.fastq | FASTQ | DNA | 830553 | 96502866 | 35 | 116.2 | 151 |
| P3-5566-I-virome_R1.fastq | FASTQ | DNA | 869113 | 99885773 | 35 | 114.9 | 151 |
| P3-5566-I-virome_R2.fastq | FASTQ | DNA | 869113 | 99970075 | 35 | 115 | 151 |
| P3-5566-Q-virome_R1.fastq | FASTQ | DNA | 895461 | 99874844 | 35 | 111.5 | 151 |
| P3-5566-Q-virome_R2.fastq | FASTQ | DNA | 895461 | 99977651 | 35 | 111.6 | 151 |
| P3-5566-Ids-virome_R1.fastq | FASTQ | DNA | 1018301 | 117549835 | 35 | 115.4 | 151 |
| P3-5566-Ids-virome_R2.fastq | FASTQ | DNA | 1018301 | 117660650 | 35 | 115.5 | 151 |
| P4-1728-I-virome_R1.fastq | FASTQ | DNA | 999725 | 113844547 | 35 | 113.9 | 151 |
| P4-1728-I-virome_R2.fastq | FASTQ | DNA | 999725 | 113966250 | 35 | 114 | 151 |
| P4-1728-Q-virome_R1.fastq | FASTQ | DNA | 969928 | 104680445 | 35 | 107.9 | 151 |
| P4-1728-Q-virome_R2.fastq | FASTQ | DNA | 969928 | 104790573 | 35 | 108 | 151 |
| P4-1728-Ids-virome_R1.fastq | FASTQ | DNA | 983342 | 114042200 | 35 | 116 | 151 |
| P4-1728-Ids-virome_R2.fastq | FASTQ | DNA | 983342 | 114146622 | 35 | 116.1 | 151 |
| P5-8986-I-virome_R1.fastq | FASTQ | DNA | 758798 | 88563713 | 35 | 116.7 | 151 |
| P5-8986-I-virome_R2.fastq | FASTQ | DNA | 758798 | 88653215 | 35 | 116.8 | 151 |
| P5-8986-Q-virome_R1.fastq | FASTQ | DNA | 891602 | 106335392 | 35 | 119.3 | 151 |
| P5-8986-Q-virome_R2.fastq | FASTQ | DNA | 891602 | 106413799 | 35 | 119.4 | 151 |
| P5-8986-Ids-virome_R1.fastq | FASTQ | DNA | 903287 | 104366876 | 35 | 115.5 | 151 |
| P5-8986-Ids-virome_R2.fastq | FASTQ | DNA | 903287 | 104467635 | 35 | 115.7 | 151 |
| P6-8992-I-virome_R1.fastq | FASTQ | DNA | 873992 | 99076446 | 35 | 113.4 | 151 |
| P6-8992-I-virome_R2.fastq | FASTQ | DNA | 873992 | 99149221 | 35 | 113.4 | 151 |

| | | | | | | | |
|---|---|---|---|---|---|---|---|
| P6-8992-Q-virome_R1.fastq | FASTQ | DNA | 746119 | 86020732 | 35 | 115.3 | 151 |
| P6-8992-Q-virome_R2.fastq | FASTQ | DNA | 746119 | 86107475 | 35 | 115.4 | 151 |
| P6-8992-Ids-virome_R1.fastq | FASTQ | DNA | 1103322 | 124352570 | 35 | 112.7 | 151 |
| P6-8992-Ids-virome_R2.fastq | FASTQ | DNA | 1103322 | 124414225 | 35 | 112.8 | 151 |

Supplementary Table 2

| | Raw PE reads | PE reads after QC filtering | Human PE reads removed | Input of total PE reads for SeqScreen | Total PE reads assigned by SeqScreen | PE reads assigned to virus | PE reads assigned to bacteria | "Dark matter" PE reads (unassigned) |
|---|---|---|---|---|---|---|---|---|
| **Viral enrichment: A=Protocol A; A.0.1=ZymoBIOMICS DNA/RNA Mini kit; A.0.2=AllPrep Power Viral DNA/RNA kit** | | | | | | | | |
| A_1 | 1664460 | 1417212 | 995 | 1416217 | 1332512 | 308 | 1278526 | 83705 |
| A.0.2_1 | 1742277 | 1419592 | 3481 | 1416111 | 1146472 | 2261 | 1075802 | 269639 |
| A.0.1_1 | 1456005 | 1181383 | 175 | 1181208 | 1050689 | 1001 | 1001584 | 130519 |
| A_2 | 1691399 | 1447463 | 2336 | 1445127 | 560427 | 5030 | 400458 | 884700 |
| A.0.2_2 | 1460188 | 1261848 | 96 | 1261752 | 1172462 | 383 | 1137410 | 89290 |
| A.0.1_2 | 1697036 | 1510265 | 76 | 1510189 | 1462573 | 340 | 1429268 | 47616 |
| A_3 | 1600505 | 1213434 | 17666 | 1195768 | 549756 | 15253 | 370831 | 646012 |
| A.0.2_3 | 1542093 | 1396271 | 91 | 1396180 | 1299294 | 523 | 1226402 | 96886 |
| A.0.1_3 | 1375155 | 1191969 | 1565 | 1190404 | 1066577 | 998 | 1011143 | 123827 |
| A_4 | 1538372 | 1329715 | 3111 | 1326604 | 355924 | 2367 | 132785 | 970680 |
| A.0.2_4 | 1499517 | 1297634 | 1343 | 1296291 | 1219353 | 354 | 1190389 | 76938 |
| A.0.1_4 | 1862747 | 1591820 | 2930 | 1588890 | 1514721 | 190 | 1480043 | 74169 |
| A_5 | 1555191 | 1356565 | 322 | 1356243 | 337225 | 1867 | 185219 | 1019018 |
| A.0.2_5 | 1673012 | 1367067 | 255 | 1366812 | 1211687 | 416 | 1092626 | 155125 |
| A.0.1_5 | 1410253 | 1237346 | 767 | 1236579 | 1171627 | 210 | 1132186 | 64952 |
| **Viral enrichment strategy: A=Protocol A (ultracentrifugation); A.1=PEG-6000** | | | | | | | | |
| A_6 | 365571 | 331725 | 12 | 331713 | 220968 | 151549 | 40337 | 110745 |
| A_7 | 368602 | 337085 | 11 | 337074 | 228147 | 155972 | 42388 | 108927 |
| A.1_6 | 307751 | 283223 | 112 | 283111 | 171909 | 106236 | 35154 | 111202 |
| A.1_7 | 321036 | 293170 | 18 | 293152 | 183264 | 118754 | 35936 | 109888 |
| **Amplification strategy: A=Protocol A (PCR); A.2=MDA** | | | | | | | | |
| A_6 | 365571 | 331725 | 12 | 331713 | 220968 | 151549 | 40337 | 110745 |
| A_7 | 368602 | 337085 | 11 | 337074 | 228147 | 155972 | 42388 | 108927 |
| A.2_6 | 227262 | 213851 | 4 | 213847 | 125069 | 40464 | 45931 | 88778 |
| A.2_7 | 260881 | 244483 | 11 | 244472 | 145078 | 49746 | 52059 | 99394 |
| **Library kit: A=Protocol A (DNA Prep library kit); A.3=TruSeq Nano DNA library Prep*** | | | | | | | | |
| A_8 | 5131783 | 4617437 | 3105 | 4614332 | 2094852 | 495400 | 999600 | 2519480 |
| A.3_8 | 1654933 | 1624596 | 1002 | 1623594 | 928508 | 181151 | 509456 | 695086 |
| A_9 | 4781559 | 4434669 | 2810 | 4431859 | 1854702 | 471774 | 728429 | 2577157 |
| A.3_9 | 1217574 | 1021570 | 685 | 1020885 | 492365 | 79663 | 229450 | 528520 |
| A_10 | 4913300 | 4540936 | 316 | 4540620 | 1620160 | 29206 | 918753 | 2920460 |

Supplementary Table 2

| | | | | | | | | |
|---|---|---|---|---|---|---|---|---|
| A.3_10 | 1694898 | 1639087 | 112 | 1638975 | 686407 | 22902 | 405360 | 952568 |
| A_11 | 3608775 | 3218914 | 3015 | 3215899 | 1335293 | 20957 | 877524 | 1880606 |
| A.3_11 | 1390669 | 1333453 | 335 | 1333118 | 707595 | 14235 | 413026 | 625523 |
| A_12 | 3811556 | 3541076 | 423 | 3540653 | 1315079 | 7922 | 660984 | 2225574 |
| A.3_12 | 1350120 | 1321047 | 113 | 1320934 | 509177 | 9196 | 278119 | 811757 |
| **DNA/RNA extraction kit: A=Protocol A (QIAamp Viral RNA mini kit); A.4=Purelink kit RNA/DNA mini kit; A.4.1=Purelink kit RNA/DNA mini kit and cDNA second strand synthesis** | | | | | | | | |
| A.4_13 | 897795 | 739396 | 39988 | 699408 | 489140 | 11477 | 368323 | 210268 |
| A_13 | 1020060 | 856378 | 3949 | 852429 | 710158 | 2043 | 664112 | 142271 |
| A.4.1_13 | 651458 | 489337 | 48254 | 441083 | 321944 | 7510 | 244292 | 119139 |
| A.4_14 | 919576 | 834110 | 2049 | 832061 | 384576 | 132099 | 124450 | 447485 |
| A_14 | 1018526 | 904308 | 414 | 903894 | 370855 | 60933 | 172416 | 533039 |
| A.4.1_14 | 830553 | 744404 | 4126 | 740278 | 359900 | 117076 | 119299 | 380378 |
| A.4_15 | 869113 | 791244 | 924 | 790320 | 442132 | 287263 | 71743 | 348188 |
| A_15 | 895461 | 804130 | 770 | 803360 | 442276 | 286729 | 71459 | 361084 |
| A.4.1_15 | 1018301 | 916744 | 2171 | 914573 | 542393 | 367658 | 79226 | 372180 |
| A.5_16 | 999725 | 908923 | 786 | 908137 | 551807 | 195550 | 212165 | 356330 |
| A_16 | 969928 | 880027 | 950 | 879077 | 510464 | 176568 | 205860 | 368613 |
| A.4.1_16 | 983342 | 906062 | 1385 | 904677 | 564773 | 205283 | 208695 | 339904 |
| A.4_17 | 758798 | 693210 | 3668 | 689542 | 249726 | 2123 | 140239 | 439816 |
| A_17 | 891602 | 821949 | 2757 | 819192 | 308578 | 2293 | 176454 | 510614 |
| A.4.1_17 | 903287 | 815061 | 11596 | 803465 | 290134 | 3451 | 163935 | 513331 |
| A.4_18 | 873992 | 789156 | 787 | 788369 | 304208 | 3569 | 129607 | 484161 |
| A_18 | 746119 | 681266 | 599 | 680667 | 277402 | 3595 | 122704 | 403265 |
| A.4.1_18 | 1103322 | 987145 | 1921 | 985224 | 384662 | 5346 | 172907 | 600562 |
| *TruSeq samples are SE reads | | | | | | | | |

### Supplementary Figures

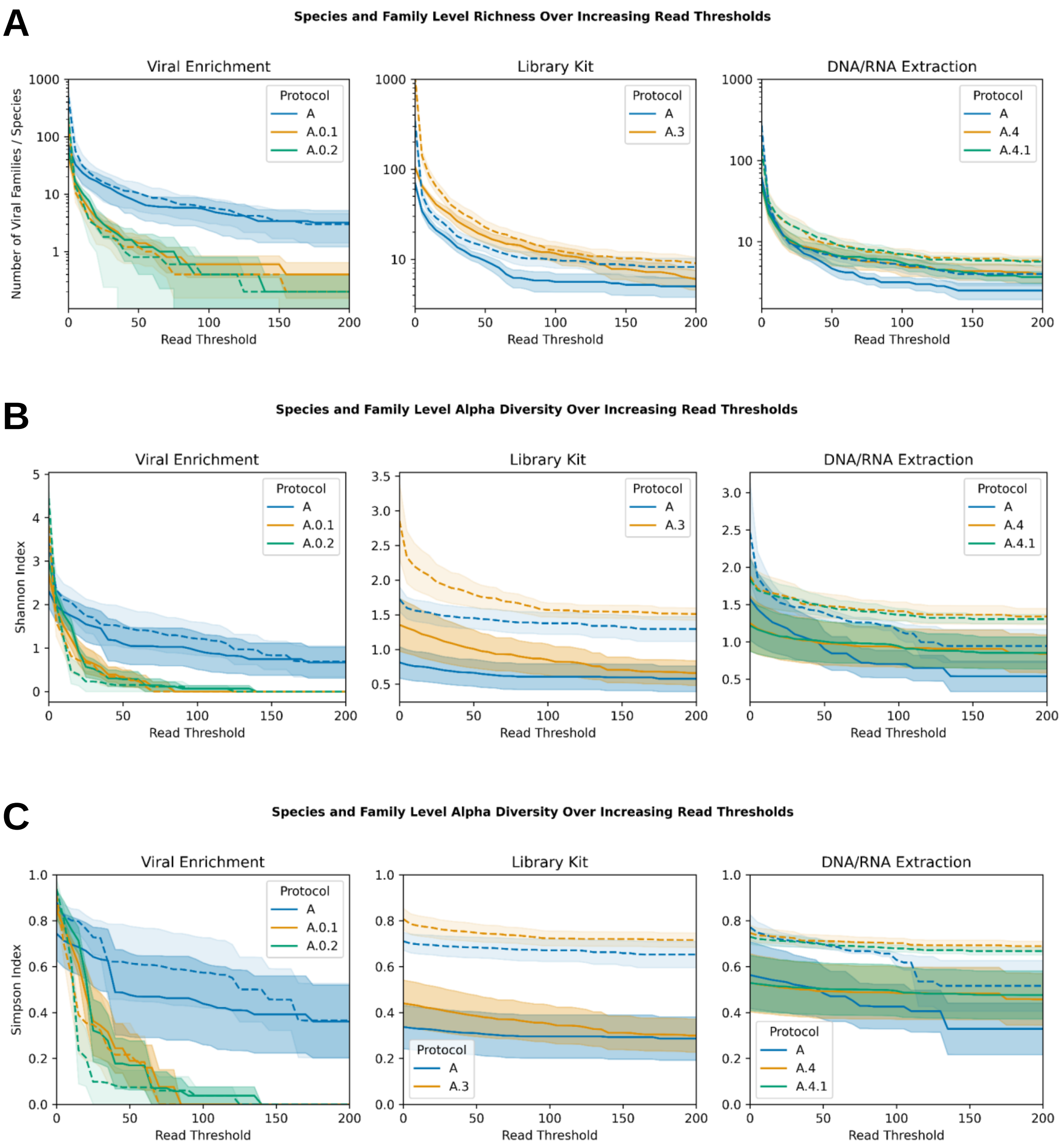

**Figure S1: Richness and diversity of viral taxonomic assignments for read thresholds below 200.** Species-level metrics are shown with dashed lines, while continuous lines represent family-level metrics. Each metric is calculated at increasing read thresholds, disregarding any taxa below the required number of reads. Cutoffs from 0-200 with step sizes of 5 were calculated. A) Species- and family-level richness. B) Species- and family-level Shannon index. C) Species- and family-level Simpson index. Protocols A.1 and A.2 were excluded from the analysis due to the small number of samples (n=2).

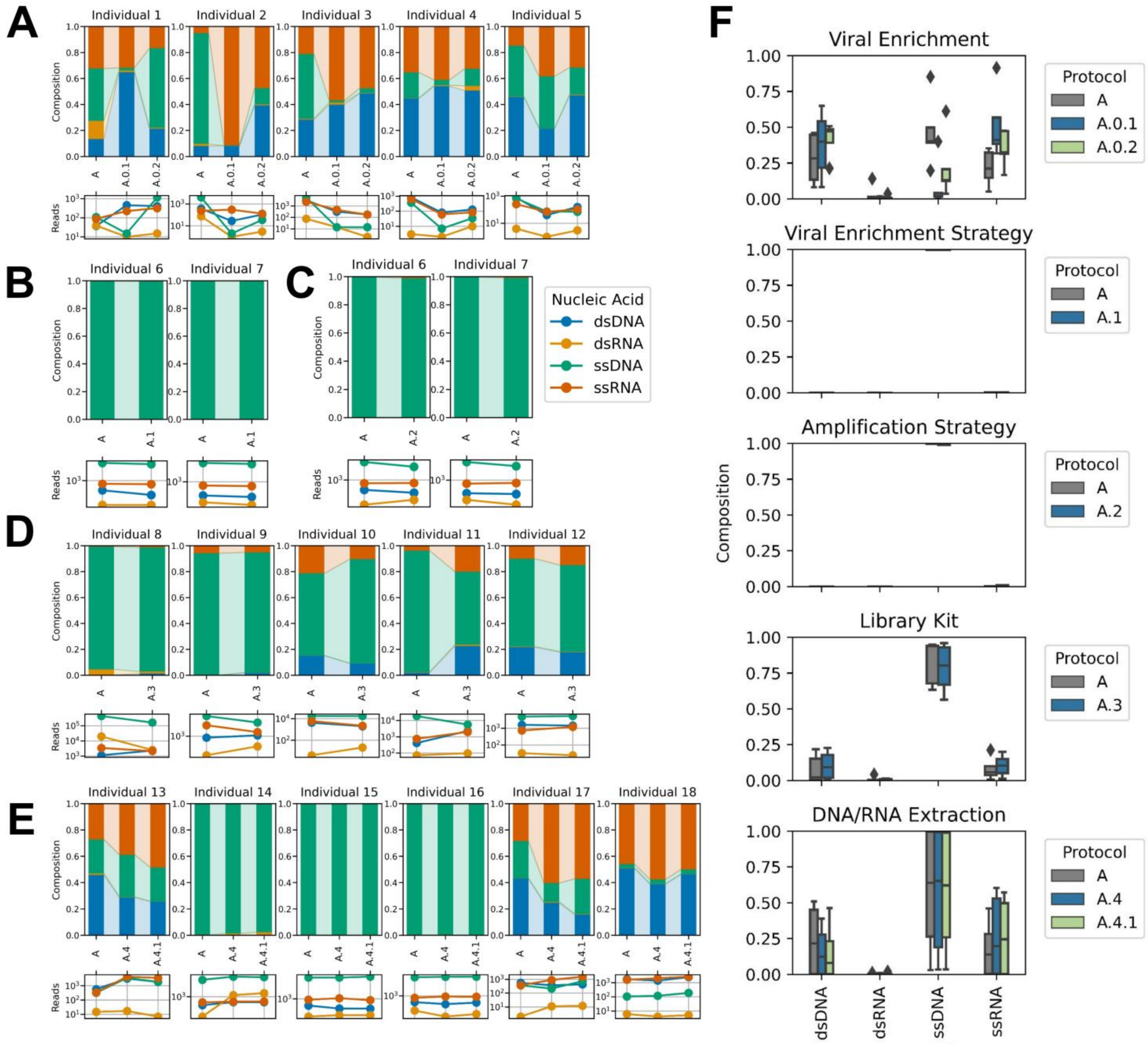

**Figure S2: Composition of viruses by genome type for the protocols tested.** For A-E, the top figures represent the genome composition of individual samples, while the bottom one show the reads of genome types (dsDNA, ssDNA, dsRNA and ssRNA). A) Viral enrichment, where protocol A corresponds to viral precipitation by ultracentrifugation and protocols A.0.1 and A.0.2 lack viral enrichment step (nucleic acids were directly extracted using ZymoBIOMICS DNA/RNA Mini kit or AllPrep Power Viral DNA/RNA kit, respectively). B) Viral enrichment strategy, where protocol A corresponds to ultracentrifugation and protocol A.1 to PEG-600-based precipitation. C) Amplification strategy, where protocol A corresponds to PCR-SISPA and protocol A.2 to MDA. D) Library preparation kit, where protocol A corresponds to the DNA Prep (M) Tagmentation kit and protocol A.3 to the TruSeq Nano DNA Library Prep kit. E) DNA/RNA extraction, where protocol A corresponds to the QIAamp Viral RNA Mini kit, protocol A.4 to the PureLink Viral RNA/DNA Mini kit and protocol A.4.1 to PureLink Viral RNA/DNA Mini kit followed by the cDNA second-strand synthesis. F) Boxplots showing the genome composition for the protocols. The central line represents the median, box limits indicate the Q1 and Q3 quartiles, and whiskers extend to Q1-1.5 IQR and Q3+1.5 IQR, marking the lowest and highest non-outlier values.

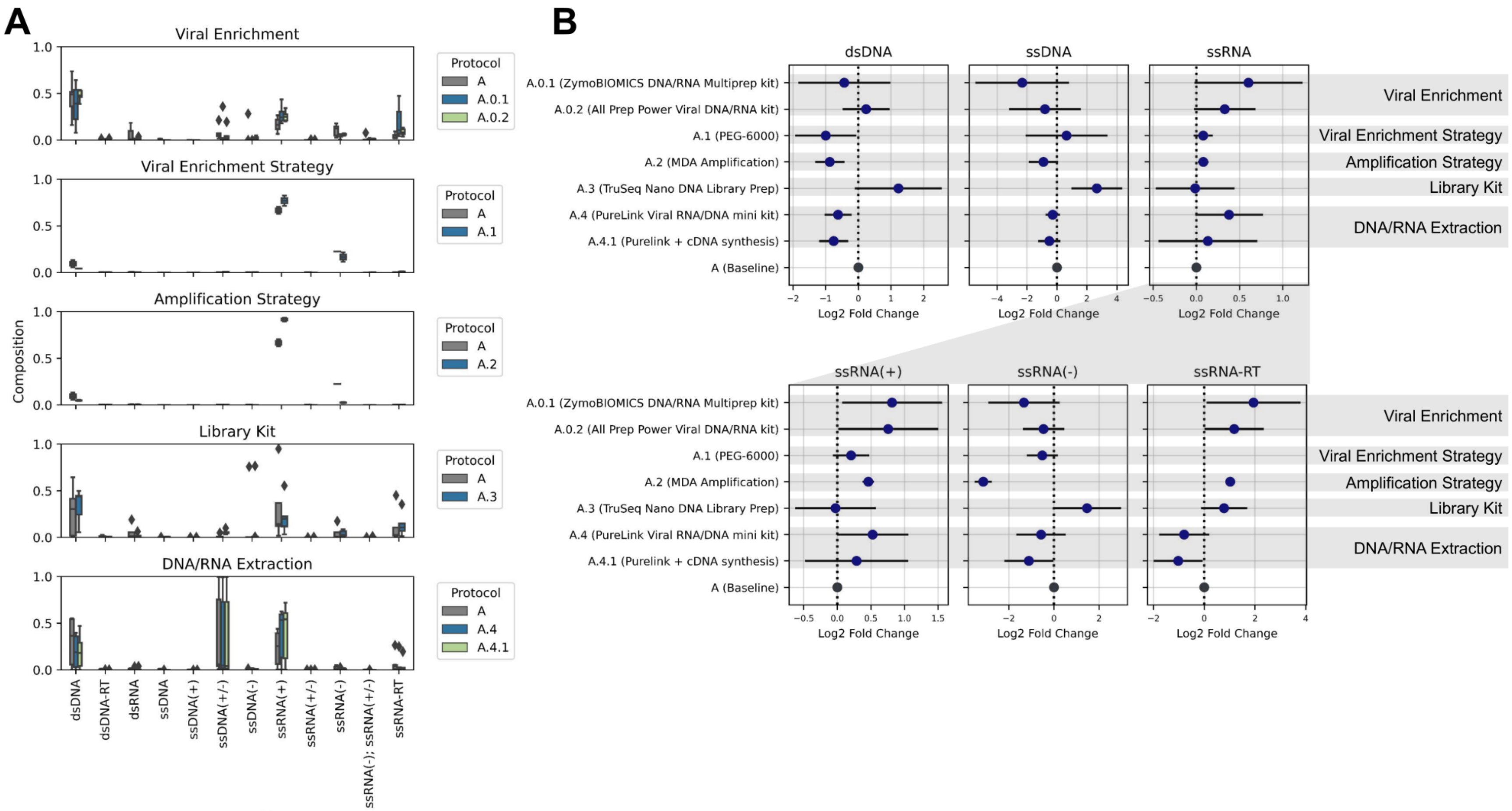

**Figure S3: Composition of viruses by genome type for the protocols tested, excluding the *Microviridae* reads.** A) Boxplots showing the genome composition excluding *Microviridae* reads. The central line represents the median, box limits indicate the Q1 and Q3 quartiles, and whiskers extend to Q1-1.5 IQR and Q3+1.5 IQR, marking the lowest and highest non-outlier values. B) Mean and 95% confidence interval (CI) of the log2 fold-change in each genome type after excluding *Microviridae* reads. Each modified protocol was compared to protocol A within its respective set of processed samples.

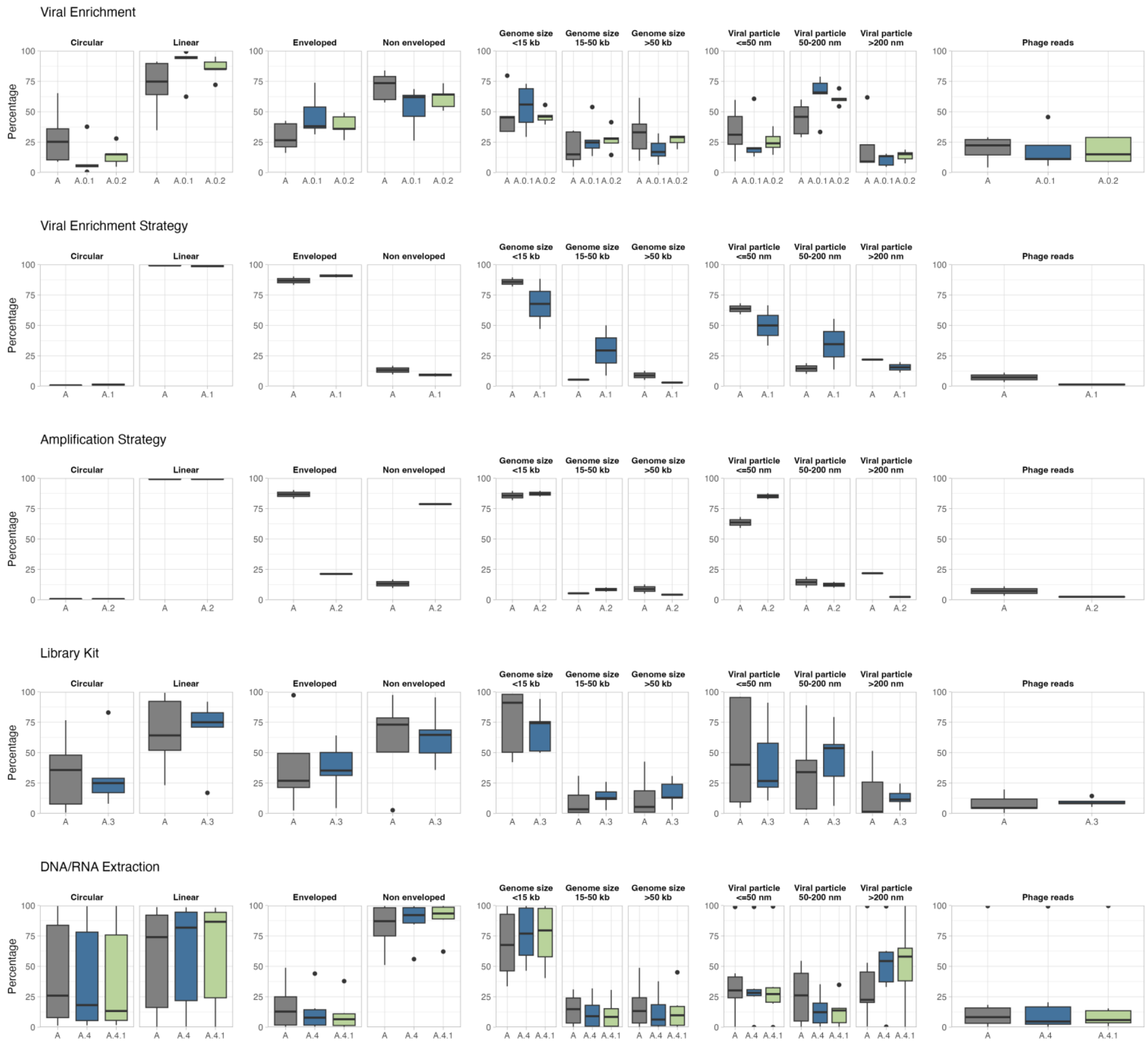

**Figure S4: Structural and genomic characteristics of viruses identified by each protocol.** Boxplots for each protocol showing the characteristics of viruses: circular or linear genome, enveloped or non enveloped structure, genome size (<15, 15-50 and >50 kb), viral particle size (<=50, 50-200 and >200 nm), and the proportion of phage-assigned particles. The central line represents the median, box limits indicate Q1 and Q3, and whiskers extend to the lowest and highest non-outlier values (Q1-1.5 IQR and Q3+1.5 IQR).
